# Genotype-Epigenome-Phenotype Integration Reveals the Contributions of Peripheral Immune Cells to Bipolar Disorder Pathogenesis, Phenotypic Heterogeneity, and Therapy

**DOI:** 10.1101/2025.03.17.25324124

**Authors:** Lei Hou, Yue Li, Xushen Xiong, Yosuke Tanigawa, Yongjin Park, Samuel W. Lenz, Amy Grayson, Jeong-Heon Lee, Euijung Ryu, Janet E. Olson, Joanna M. Biernacka, Mark A. Frye, Tamas Ordog, Manolis Kellis

## Abstract

Immune dysfunctions are believed to contribute to bipolar disorder (BD), yet their mechanistic basis remains unclear. To address this, we systematically characterize BD-associated epigenomic and genetic variation in peripheral blood immune cells by profiling and integrating 833 genome-wide maps of five histone modification marks across 180 individuals (88 Type I BD patients, 92 controls), coupled with whole-genome sequencing data and rich medical records. We annotate 450k candidate *cis*- regulatory elements (CREs) and identify differential CREs (dCREs) in BD patients, suggesting down-regulated adaptive and up-regulated innate immune response. We predict candidate BD driver genes in the circulating immune system, which frequently show matched brain activity mainly related to calcium signaling and endoplasmic reticulum (ER) transport, suggesting dysregulated synaptic transmission, neuronal plasticity, and ER stress. We find that candidate driver genes are often linked to BD GWAS variants through blood-specific eQTLs not found in any brain cell types, indicating potential causal roles of circulating immune cells in bipolar disorder. We then infer 24 latent factors of BD-differential CRE variation and use them to group the patients into five epigenomic subtypes, which also show distinct disease phenotypes, including infection and inflammation, osmotic laxative use and glucose intolerance, quetiapine use, and hypertension. We next associate immune-partitioned BD polygenic risk scores with patient epigenomic subtypes, revealing the genetic basis of BD patient heterogeneity captured by blood epigenomics. Lastly, by analyzing transcriptional responses to known pharmacological interventions in hematopoietic cells that enrich BD patient group-specific dysregulated genes, we identify drugs/compounds that could be repurposed for ameliorating BD-associated immune dysfunction in a patient group-dependent manner. Overall, based on our study of genotype-epigenome-phenotype integration, we infer a potentially causal role of immune cells in BD, offering insights into biomarkers, subtypes, and precision medicine interventions targeting peripheral immune dysfunction and thus advancing precision medicine in BD.

## Introduction

The substantial global burden of bipolar disorder (BD), schizophrenia, and other psychiatric diseases represents a significant public health concern^1^. These disorders often necessitate lifelong treatment, which entails a considerable financial burden and negatively impacts the patient quality of life. Management is challenging due to the considerable heterogeneity observed among patients^2–4^ and the temporal variability of disease course^5^. Multiple sources of heterogeneity, including genetic factors, environmental influences, and comorbidities, complicate early diagnosis, hinder the accurate prediction of medication efficacy and response, and make targeted interventions challenging, despite the availability of numerous pharmacological options. To advance our understanding and improve patient outcomes, two interdependent efforts are essential: (1) elucidating the diverse genetic and non-genetic factors that underlie pathogenesis and drug response and (2) identifying the molecular and genetic signatures of different disease subtypes that may be predictive of distinct physiological states and drug response.

Recent genome-wide association studies (GWAS) have revealed tens to hundreds of loci associated with different psychiatric disorders, including bipolar disorder (BD)^6^, schizophrenia (SCZ)^7^, and major depressive disorder (MDD)^8^. As expected, top genetic hits for BD implicate genes that directly impact neuronal activity, including ion channels, synapse function, and neurotransmitters. These are also the primary targets of BD medicines, for example, lithium modulating neurotransmitter release^9^, valproate inhibiting voltage-gated sodium channels and modulating glutamate and calcium channels^10^, and quetiapine antagonizing dopamine D_2_ and serotonin 5-HT_2A_ receptors^11^. GWAS, an unbiased analytical tool, has also revealed that immune function genes may be important causal contributors to psychiatric disorders^12–14^, with driver genetic loci associated with multiple psychiatric disorders enriched in immune-related pathways and genomic regions active in immune cell types^15,16^. These disorders also show a strong genetic correlation with inflammatory or autoimmune diseases and inflammatory biomarkers in blood, such as C-reactive protein^17^. Based on cell biological evidence, microglia, a resident immune cell type in the brain, is considered central for brain homeostasis as their dysregulation may lead to neuroinflammation^18^, a contributing factor to psychiatric diseases^19^. Beyond the immune system in the brain, there is also accumulating evidence supporting the role of circulating immune cells in psychiatric diseases, which can both impact and be affected by brain activity through bidirectional communication with neurons via cytokines and chemokines^20^. Multiple studies have reported alterations of immune functions in peripheral blood from BD, SCZ, and MDD patients, including T cell exhaustion and T cell senescence^21,22^. Common BD medicines, e.g., aripiprazole, topiramate, and quetiapine, could lower inflammation in peripheral blood, indicating a potential mechanism of action^23^. Immunomodulatory agents, such as non-steroidal anti-inflammatory drugs (NSAIDs) and cytokine inhibitors have also shown efficacy in subsets of MDD or BD patients with high baseline inflammation or a history of childhood adversity^24^, which is known to affect immune functions^25^.

We hypothesized that circulating immune cells may not merely act as effectors of other etiological factors of BD, but also directly impact the pathogenesis, especially in subgroups of patients. We reasoned that epigenomic profiling could both provide tissue/cell type-specific regulatory activity for GWAS hits—most of which are non-coding variants—and capture inter-individual variations stemming from genetic and other risk factors, such as health conditions, serving as an ideal approach to test our hypothesis. We generated 833 epigenetic profiles for five histone modifications in peripheral blood buffy coat samples from 180 participants including 88 type I BD patients (i.e., BD patients with manic and, usually, depressive episodes) and 92 age- and gender-matched controls without any psychiatric condition or disorder. We took five steps to analyze epigenomic variation by integrating these datasets with disease status, genetic variants from whole-genome sequencing (WGS) data, and risk factors and other health conditions from electronic health record (EHR) data across 3286 phenotypic variables for BD patients^26^ **(Fig. 1a)**. We first identified 803-22k *cis*-regulatory elements (CREs) across the five histone marks showing differential activity between BD patients and controls (differential CREs, dCREs), suggesting overall increased activity of innate immune response and repression of adaptive immune response in BD. We inferred 2632 genes with BD-differential epigenomic signatures based on dCREs at gene body and gene regulatory domain defined by promoter capture Hi-C data. We next tracked the impact of BD-associated genetic variants based on GWAS data and inferred driver genes in immune cells. We mapped 1.8k-27k genetically influenced CREs (gCREs) implicating 1.6 M histone quantitative trait loci (hQTLs) in blood and identified 341 gCREs showing BD–GWAS–hQTL colocalization, which were enriched in genes related to calcium signaling and endoplasmic reticulum processes. We then predicted 215 candidate BD driver genes acting in circulating immune cells based on BD GWAS signals, QTLs affecting CRE activity, CRE–gene links, and BD-differential epigenomic signatures. Of these 215 genes, 28 were confirmed by GWAS–eQTL colocalization in white blood cells only, whereas 11 were confirmed in both brain and blood cell types, suggesting an immune component in BD pathogenesis. We also applied latent variable analysis to capture major epigenomic variation across individuals, mapped patients into a lower-dimension space, and clustered them into five subgroups, with enriched clinical terms indicating BD comorbidity, risk factors, and preferred medicines. We found that this patient subtypes were significally associated with several BD polygenic risk scores (PRSs), especially PRSs partitioned by BD genetics-associated gCREs. Finally, drug repurposing analysis based on the enrichment of BD dysregulated genes in differential gene expression profiles in response to pharmacological perturbations identified 428 drugs and compounds, including drugs targeting tumors, the nervous system, and the cardiovascular system, as potential treatment options for BD.

**Figure 1.**
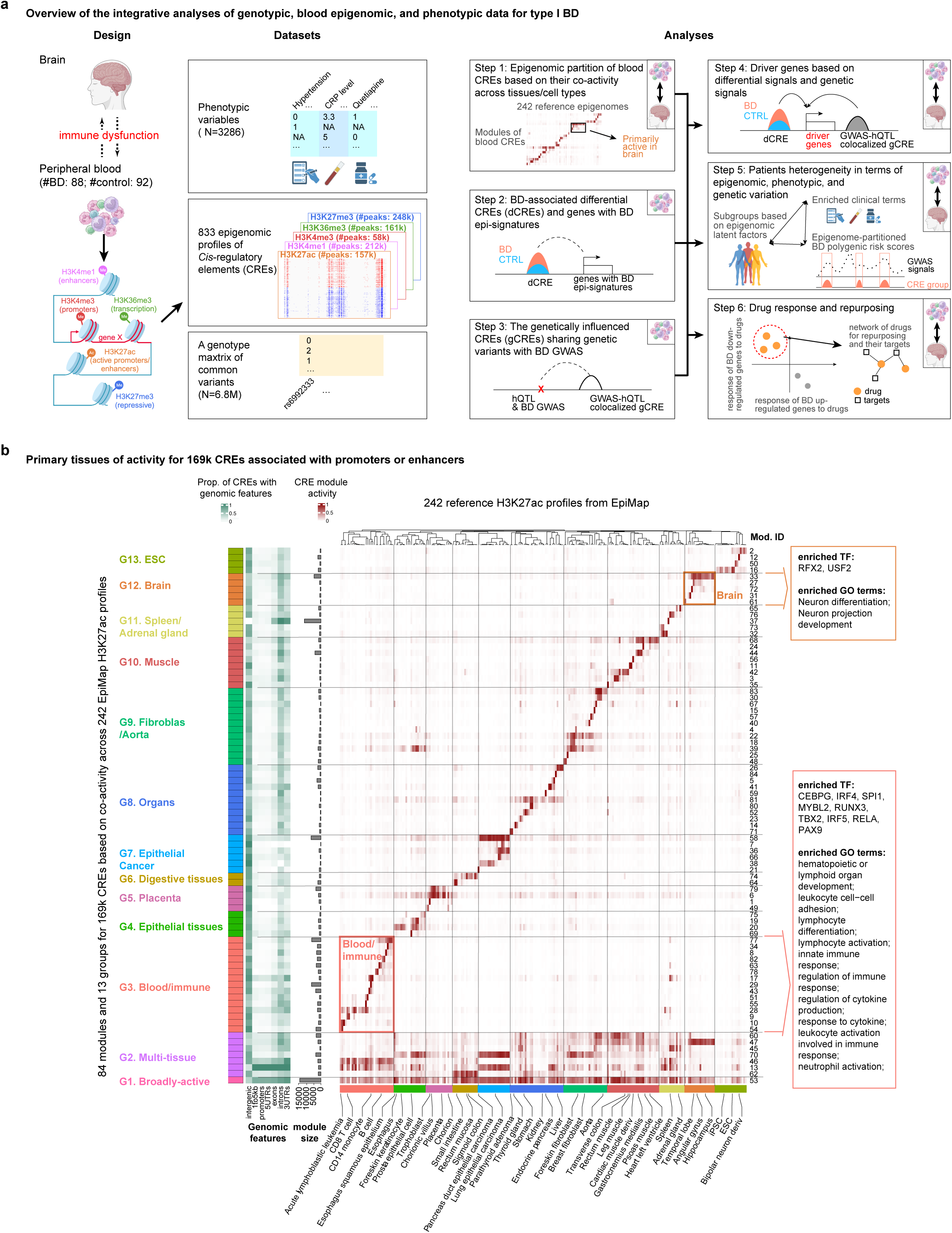
Overview of the study and epigenomic profiling. **a,** Overview of the integrative analyses of genotypic, blood epigenomic, and phenotypic data for type I bipolar disorder (BD). Panels from left to right: study design to reflect the connection between brain and blood; data generated including blood epigenomic, genotypic, and phenotypic data; and the integrative analyses consisting of five steps. **b**, 169k CREs associated with enhancers or promoters (H3K27ac, H3K4me1, and H3K4me3) in this study forming 13 groups of 84 modules based on coactivity across 242 reference epigenomes. Panels from top to bottom: 84 CRE modules; proportion of CREs annotated in different genomic features for each module; size of each module; average CRE activity (H3K27ac signal) of each module (by column) across 242 reference epigenomes (by row). Representative sample names labeled for each sample cluster on the right; IDs of 84 CRE modules labeled at the bottom.

In summary, we present the largest blood histone modification profiling dataset in BD to date and comprehensive integration analyses that reveal the role of circulating immune components in BD based on genetic signals, epigenomic marks, and gene expression. We also suggest drugs and targets based on BD-associated immune dysfunction. Our study demonstrates the important role of white blood cells, a readily accessible biospecimen, in understanding BD pathogenesis and patient heterogeneity.

## Results

### An epigenomic atlas of 439k blood CREs across 180 individuals

To capture epigenomic differences in BD patients compared to control individuals, we profiled the genome-wide distribution of five histone marks using chromatin immunoprecipitation followed by high-throughput sequencing (ChIP-seq) in blood buffy coat samples from 180 participants including 88 patients and 92 controls from Mayo Clinic, from mainly non-Hispanic white population(**Fig. 1a**). These five widely studied marks relevant to gene-regulatory processes include H3K27ac, which is associated with active promoters and enhancers, H3K4me1, a marker of enhancer regions, H3K4me3, a marker of promoters, H3K36me3, a mark that reflects transcriptional elongation, and H3K27me3, which is associated with Polycomb-repressed regions^27^.

Across all subjects, we identified 157k *cis*-regulatory elements (CREs) with peaks in the epigenomic profiles of H3K27ac, 212k CREs for H3K4me1, 58k for H3K4me3, 161k for H3K36me3, and 248k CREs for H3K27me3 (**Fig. 1a, Supplementary Table 2**), covering 439k non-overlapping regions in the genome. Distribution analysis of CREs around genes showed that 76-88% of CREs across the five histone marks were located within 50 kb of the transcription start sites (TSS) or transcription termination sites (TTS) of a gene (**Extended Data Fig. 1a**). Consistent with being a mark of transcriptional elongation^28^, H3K36me3 CREs showed much higher average signals within gene bodies compared to promoter regions than other CREs (0.81 vs. 0.18-0.35). The observed overlaps among different types of CREs were also consistent with their functions: We found 82% of H3K27ac CREs (active enhancers and promoters) overlapping with H3K4me1 CREs (active or poised enhancers), 84% of H3K4me3 CREs (active and poised promoters) overlapping with H3K27ac CREs, but only 3% of H3K27me3 CREs (repressed genomic regions) overlapping with H3K36me CREs (transcribed regions) (**Extended Data Fig. 1b**).

As CREs are frequently shared between tissues, we next sought to understand the activity profiles of these blood CREs in a broad collection of tissues. Specifically, we quantified the activity of 169k CREs annotated by the enhancer/promoter-associated marks H3K27ac, H3K4me1, and H3K4me3 based on the H3K27ac signal across 242 reference epigenomes from the Encyclopedia of DNA Elements (ENCODE), the National Institutes of Health Roadmap Epigenomics Mapping Consortium database, and the National Human Genome Research Institute’s Genomics of Gene Regulation (GGR) project^29–31^ and integrated in EpiMap^32^. Based on these profiles, we clustered them into 84 modules and 13 groups (G1-G13, **Fig. 1b**). We found these blood CREs to be broadly active (G1) or active in multiple tissues (G2), with the rest being active predominantly in immune/blood cells (G3), epithelial/epidermal cells (G4), the placenta (G5), digestive tissues (G6), epithelial cancer (G7), miscellaneous organs (G8), fibroblast/aorta (G9), muscle (G10), spleen/adrenal (G11), brain (G12) and embryonic stem cells (G13).

We then investigated the enrichment of transcription factor (TF) binding motifs and gene ontology (GO) terms for each CRE module. The CREs broadly active or active in multiple types of tissues (G1 and G2) were more likely to appear in promoter regions (40% vs. 12% in the background), while those active only in certain tissue types were more likely to be from intergenic regions (31% vs. 26% in the background) **(Fig. 1b)**, consistent with the previous findings that tissue specificity is mostly driven by intergenic enhancers^29^. By TF motif and GO enrichment analysis, we found immune-related TFs, such as IRF4^33^, SPI1^34^, and IRF5^35^, and immune response-related biological processes enriched in G3 (**Extended Data Fig. 1c-e**), confirming the functions of CREs primarily active in blood or immune cells. Interestingly, we also found CREs that were predominantly active in the brain (G12). This group was also enriched in two neuron-related TFs, RFX2^36^ and USF2^37^, and neuron differentiation-related GO terms, indicating potential sharing between blood and brain.

### Differential activity of CREs in bipolar disorder

To explore the role of immune components in BD, we first asked whether the activity of CREs in blood is different in BD patients compared to controls. Since the buffy coat samples are mixtures of different immune cell types, the variation of CRE activity may also be impacted by cell fraction variation. We therefore estimated for each patient the cell fractions of six major white blood cell types, including CD4^+^ T cells, CD8^+^ T cells, B cells, natural killer cells, monocytes, and neutrophils, with our deconvolution approach (**Methods**). Neutrophils showed the largest fraction, and these estimated fraction varied across individuals (**Extended Data Fig. 2a**). We next identified differential CREs (dCREs) between BD patients and controls for each histone mark. We applied surrogate variable analysis followed by a linear regression model to account for the hidden variables (**Methods)**. As expected, inferred surrogate variables from different histone modifications were strongly correlated with the estimated cell fractions (see H3K27ac as an example in **Extended Data Fig. 2b**), suggesting that the impact of cell fraction variations was captured and thus corrected during dCREs analysis.

In total, we detected 22k (H3K27ac), 16.1k (H3K36me3), 1.6k (H3K27me3), 805 (H3K4me1), and 1.6k (H3K4me3) dCREs (adjust *P* 0.05 based on), showing the largest number of overlapped dCREs between the active marks, such as the up-regulated dCREs shared between H3K36me3 and H3K27ac (N=1638), and the down-regulated dCREs shared between H3K4me3 and H3K27ac (N=576) (**Extended Data Fig. 2c, Supplementary Table 3**). GO enrichment analysis by GREAT^38^ also confirmed coordinated impacts of BD on CREs across different marks. The repressive regulation in BD (up-regulated repressive marks and down-regulated active marks) was enriched for gene silencing and adaptive immune response, while the active regulation in BD (up-regulated active marks and down-regulated repressive marks) was enriched for neutrophil activation (**Fig. 2a**). These results confirm an elevated level of inflammation and reduced adaptive immune response in BD^39^ and highlight the utility of epigenomic analysis for capturing altered physiological states in blood. Interestingly, several neuron-related pathways were also affected in peripheral immune cells in BD, including *detection of chemical stimulus involved in sensory perception of smell*, *positive regulation of synaptic transmission, cholinergic*, and *neuron fate commitment*. This suggests shared molecular machinery between immune cells and neurons in BD.

**Figure 2.**
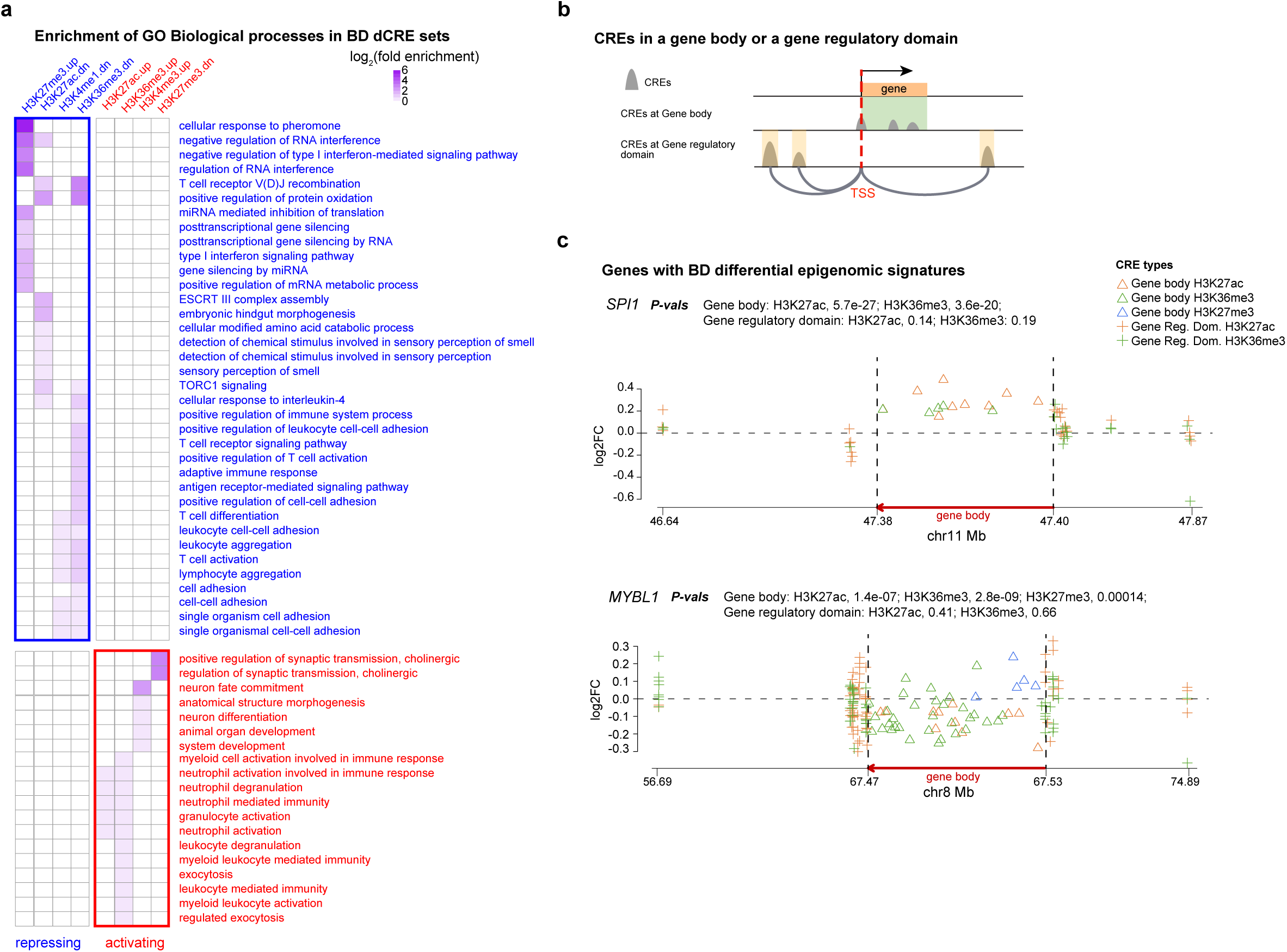
dCREs in BD and genes with BD-differential epigenomic signatures. **a,** Enrichment of GO Biological processes in BD dCRE sets. The heatmap shows the log_2_ fold enrichment of GO terms (by row) for BD dCRE sets (by column) for each histone mark with different signs, where up-regulation of repressive marks and down-regulation of activating marks are grouped as “repressing” in blue and down-regulation of repressive marks and up-regulation of activating marks are grouped as “activating” in red. b, Schematic diagram to show CREs in a gene body or in a gene regulatory domain. **c,** Two examples of genes with BD-differential epigenomic signatures. *SPI1*: consistently up-regulated dCREs in the gene body based on active marks (H3K36me3 and H3K27ac): *MYBL1*: consistently down-regulated dCREs based on active marks (H3K36me3 and H3K27ac) and up-regulated dCREs based on repressive marks (H3K27me3) in the gene body (scaled to be ⅓ of the x-axis).

We next investigated TFs that may underlie these epigenomic alterations in BD by using the motif enrichment analysis software Homer^40^. We identified 19 TF families showing motif enrichment in any dCRE set (**Supplementary Table S3**). Of the representative TFs, 15 were associated with BD based on GWAS signals, differential signals, or mechanistic evidence. Six of these TFs (IRF8, SPI1, CEBPB, RORA, ATF1, and EGR3) are directly related to inflammatory response, while five TFs (SPI1, MECP2, MBD2, JDP2, and CTCF) function as chromatin regulators/remodelers. 18 of these dCRE-associated TFs also formed a protein-protein interaction network indicating potential regulatory mechanisms connecting inflammatory responses to chromatin remodeling in the peripheral immune system of BD patients (**Extended Data Fig. 2d**).

We next inferred differential signals at the gene level. We generated multiple differential scores by meta-analysis of differential signals of CREs from each histone mark using two CRE–gene mapping strategies: (1) CREs located within the gene body and (2) CREs located within the gene regulatory domain defined as the genomic regions linked to the promoter of the gene based on promoter capture Hi-C data in immune cells (**Fig. 2b, Methods**). H3K36me3 at the gene body is an epigenomic mark for transcription elongation^41^ and is thus a reasonable surrogate for transcriptional activity. We found that two scores from the gene body (H3K27ac and H3K27me3) and two scores from the gene regulatory domains (H3K27ac and H3K36me3) were strongly correlated with the score for gene body H3K36me3 (**Extended Data Fig. 2e-f**). Our findings are also consistent with the known association between these three marks (H3K27ac, H3K36me3, and H3K27me3) and the level of gene transcription^42^. We then aggregated these five strongly correlated scores and inferred the genes with BD down-regulated (N=631) and up-regulated (N=2001) epigenomic signatures (**Extended Data Fig. 2g, Supplementary Table 3, Methods**). GO enrichment analysis of these genes with Enrichr showed downregulated adaptive immune response and up-regulated innate immune response (**Supplementary Table S3**), consistent with the results for BD dCREs above (**Fig. 2a**). The genes with BD-differential epi-signatures included six of the TFs we earlier identified as potential regulators for the dCREs (SPI1, FLI1, MYBL1, MBD2, JDP2, and CEBPB) (**Extended Data Fig. 2d**), genes associated with neutrophil extracellular traps (TLR4 and PADI4)^43^, and differentially expressed genes from previous BD blood studies^44,45^ that are involved in DNA damage (OGG1) and the TNF-α signaling pathway (TNFAIP6, TNFRSF10C, and TNFAIP2). Notably, these genes also showed consistent changes in peripheral immune cells in association with lower back pain^46^ (**Extended Data Fig. 2h)**, suggesting a shared immune component between lower back pain and bipolar disorder.

### Genetic regulation of CRE activity and implication of CRE activity in BD genetic associations

To dissect BD-associated genetic variants, we next sought to identify the impacts of genetic variants on CRE activity by histone quantitative trait locus (hQTLs) mapping. We followed the GTEx eQTL mapping protocol as follows^47^: (1) identify peer factors, (2) include top peer factors as covariates together with age and gender during hQTL mapping, and (3) correct for multiple testing based on permutation. We also verified that the cell fraction variation was corrected for during QTL mapping by confirming that the top peer factors were strongly correlated with the cell fractions estimated above (**Extended Data Fig. 3a**). We finally identified 1.5M H3K27ac hQTLs (targeting 17k genetically influenced CREs, or gCREs), 2.2M H3K4me1 hQTLs (27k gCREs), 200k H3K4me3 hQTLs (1.8k gCREs), 1.2M H3K36me3 hQTLs (13k gCREs), and 1.4M H3K27me3 hQTLs (19k gCREs) (**Extended Data Fig. 3b, Supplementary Table 4, Methods**). As expected, a major proportion of the lead hQTLs (40-70% across the five histone marks) were within 20 kb of the center of the target peaks, showing significant enrichment compared to the background (Kolmogorov–Smirnov test *P*=2.2×10^-16^) (H3K27ac shown as an example, **Extended Data Fig. 3c**).

**Figure 3.**
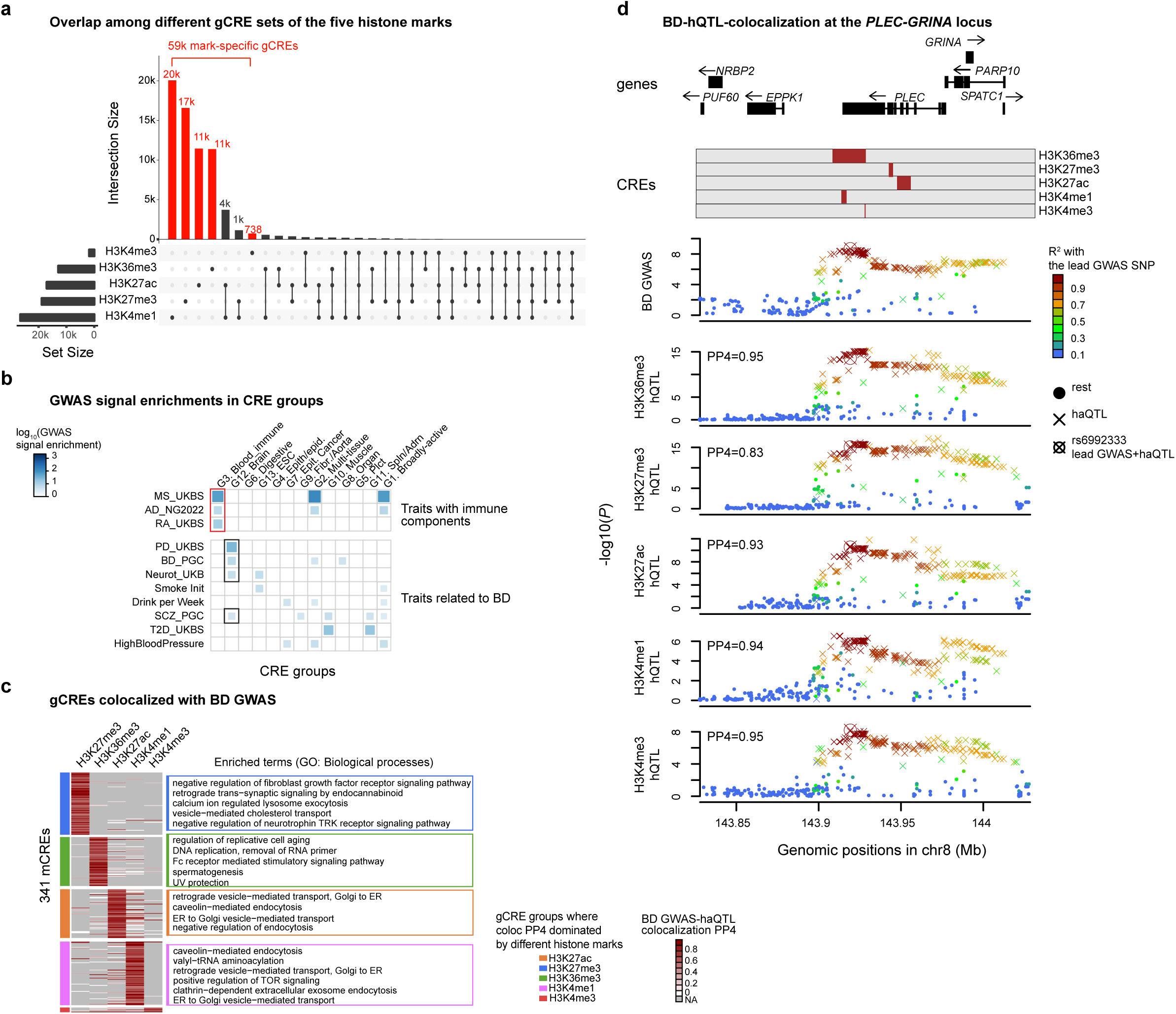
Genetics of CREs in blood and implications in BD. **a,** Overlap among different gCRE sets of the five histone marks. The upset plot shows the number (y-axis) of CREs with various combinations of histone marks, with mark-specific CREs highlighted in red. **b,** GWAS signal enrichments in CRE groups. The heatmap shows the log_10_ enrichment of different GWAS signals (by row, with groups labeled on the left) in different groups of CREs defined in Fig. 1b (by row). **c,** gCREs colocalized with BD GWAS. The heatmap shows the BD GWAS–hQTL colocalization signals for each merged CRE (mCREs) across various marks, with enriched biological processes annotated on the right. **d,** BD–hQTL-colocalization at the *PLEC-GRINA* locus shown as an example of the colocalization of BD GWAS signals and hQTL signals across different marks with the genomic positions of genes and CREs annotated on top.

Given the substantial overlap among CREs labeled by different histone marks (**Fig. 1b**), we next investigated the likelihood of their shared genetic regulation. We found that the majority of gCREs were specific to individual marks, 59k in total across the genome, demonstrating the value of our multi-mark approach to capture genetically influenced regulatory elements (**Fig. 3a**). We also identified 4k gCREs shared between H3K27ac and H3K4me1, and 1k gCREs shared between H3K27me3 and H3K4me1 (**Fig. 3a**). Following our previous studies on QTL sharing across tissues based on directionality consistency^48^, we revealed pairs of histone marks with higher degree of gCRE sharing (e.g., among H3K27ac, H3K4me1, and H3K4me3), consistent with their active regulatory roles, and others with low sharing levels (e.g., between H3K36me3 vs. H3K4me1, and H3K4me3 vs. H3K27me3), indicating complex genetic regulations across different histone marks (**Extended Data Fig. 3d**).

Based on the different annotations of CREs from blood (CRE groups across various primary tissues/cell type in **Fig. 1b**, blood dCREs in BD in **Fig. 2a**, and gCREs in **Fig. 3a**), we next asked whether the annotations could shed light on the contexts of genetic variants associated with complex diseases/traits with known immune components or related to bipolar disorder. We carried out stratified LD score regression (LDSC) analysis^49^ to quantify the enrichment of GWAS signals overlapping with different CRE annotations. Immune-related diseases, including multiple sclerosis (MS), rheumatoid arthritis (RA), and Alzheimer’s disease (AD), showed strong enrichment of GWAS signals in CREs predominantly active in blood/immune system (G3) and in gCREs (H3K27ac or H3K4me1), confirming the strong immune components in these diseases (red boxes in **Fig. 3b** and **Extended Data Fig. 3e**). Interestingly, these diseases also showed significant enrichment of GWAS signals in BD dCREs (H3K27ac, H3K36me3, and H3K4me1), indicating they might share immune components with BD. BD, together with Parkinson’s Disease (PD), neuroticism, and schizophrenia showed significant GWAS signal enrichment in CREs detected in blood and predominantly active in brain or in neurons (G12) (black boxes in **Fig. 3b**), indicating that CREs in the blood may mirror similar epigenetic regulation in the brain, and capture the impact of these disease-associated genetic variants.

To identify specific gCREs that share genetic drivers with BD and thus play important roles in BD etiology, we performed BD GWAS–hQTL colocalization analysis by the R package coloc^50^. We identified 341 merged BD-associated gCREs from different marks in our dataset (posterior probability, coloc PP4>0.5), the majority of which are histone-mark-specific gCREs (**Fig. 3c**). Interestingly, enrichment analysis by GREAT revealed that genes near these gCREs had convergent functions related to the endoplasmic reticulum (ER), indicating that ER homeostasis in circulating immune cells may contribute to BD etiology^51,52^ (**Fig. 3c**). In total, we identified 97 BD GWAS loci (*P*<=1 × 10^-6^) potentially regulating blood gCREs of any mark based on GWAS–hQTL colocalization (**Supplementary Table 4**). We found gCREs of a single mark could at most explain 60% of these loci (H3K4me1, N=58), demonstrating the necessity of our multi-mark approach to comprehensively dissect the GWAS signals. We also found 61% of these loci were associated with gCREs from multiple histone marks. For example, at the *PLEC*-*GRINA* locus, which has been linked to ER homeostasis^53,54^, hQTL signals across all five histone marks showed strong colocalization (posterior probability, coloc PP4 >0.8) with the BD GWAS signal (**Fig. 3d**), indicating a broad impact of BD-associated genetic variants over the epigenetic landscape. Notably, *GRINA* was previously identified as a differentially expressed gene in the brain samples of individuals with major depressive disorder^55^, a disorder closely related to BD.

In light of the BD GWAS signal enrichment in the CREs primarily active in the brain (G12. Brain, **Fig. 3b**) and the BD GWAS–hQTL colocalization events in the blood (**Fig. 3c**), we next asked whether the GWAS–hQTL colocalizations observed in the blood might mirror colocalization in the brain. Such a scenario could be supported by enrichment of BD GWAS–hQTL colocalization events for the gCREs shared between the blood and the brain. We first found that among the blood H3K27ac gCREs (N=30.0k), those shared in the brain (N=12.6k) based on our previous eGTEx study^56^ significantly enriched gCREs near BD GWAS signals (within 100kb, N=982, Fisher’s exact test *P*=0.0032). However, when we considered the subsets of the blood H3K27ac gCREs near BD GWAS signals (N=982), the gCREs shared in the brain (N=456) did not enrich the gCREs showing BD GWAS–hQTL colocalization (coloc PP4>0.5, N=103, Fisher’s exact test *P*=0.29), and neither did the gCREs sharing hQTL regulation between the blood and the brain (brain shared hQTL *P*<=0.01, N=31, shared hQTLs defined in **Extended Data Fig. 3f** and **Methods,** Fisher’s exact test *P*=0.46). Together, our results suggest that the BD GWAS variants we captured from hQTLs in the blood may specifically impact CRE function in the circulating immune cells. Alternatively, these BD GWAS variants may function in immune cells both in the blood and the brain but were only captured by blood hQTLs because of a larger power due to both a larger sample size and a much larger number of immune cells in the blood than in the brain. Either explanation indicates a potentially causal role of the circulating immune system in BD etiology.

### BD driver genes and blood-brain sharing

We next sought to prioritize potential BD driver genes in blood. We reasoned that the driver genes should both be driven by the BD GWAS signals and show BD-differential signals at their gene regulatory domains (CREs linked to their promoters). We first explored the global relationship between BD GWAS signals and BD differential signals at the level of chromatin interaction blocks, namely, clusters of chromatin regions connected by links within 1Mb based on promoter capture Hi-C data in immune cell types from BLUEPRINT^57^. For each of the 1370 chromatin interaction blocks identified, we quantified the enrichment of the dCREs for each histone mark over the background, and the density of the links connecting the chromatin regions. Our results showed positive correlation between link density and dCRE enrichment for the blocks with dCREs across all five histone marks (**Extended Data Fig. 4a**), indicating that the dCRE signals may have impacted each other through these chromatin interactions inside each block. Together with the gCREs associated with BD genetic signals based on colocalization or Mendelian randomization, analysis at the level of chromatin interaction blocks revealed that the blocks annotated with BD GWAS signals were more likely to show enrichment of dCRE signals, except H3K27me3-downregulated dCREs and H3K4me3-upregulated dCREs (**Extended Data Fig. 4b**). These results suggested that BD GWAS signals at these chromatin interaction blocks contribute to blood epigenomic dysregulation in BD, supporting that driver genes for BD could be identified in blood samples.

To identify driver genes based on BD-associated gene regulatory domains, we leveraged two types of CRE–gene linking evidence: (1) physical evidence, which is based on promoter capture Hi-C data in immune cells^57^, and (2) genetic evidence, namely, gLink scores, which we defined in our eGTEx study^56^. We assessed the consistency between the two pieces of evidence and confirmed that gLink scores outperformed background and the permuted scores (controlling for CRE–gene distances) in predicting Hi-C linkages (**Extended Data Fig. 4c, Methods**). We then prioritized driver genes as those linked to both upstream BD genetic signals—specifically gCREs associated with BD genetics (either by GWAS–QTL colocalization or Mendelian randomization)—and downstream epigenomic signals, namely BD dCREs. Using this method, we identified 161 and 91 BD driver genes in blood via CRE—gene linking based on Hi-C and gLink scores (overlap N=37), respectively. Consistent with the result at the level of chromatin interaction blocks, genes linked to dCREs showed enrichment in genes with BD GWAS-associated regulatory elements from the two linking strategies (*P*=1.3 × 10^-38^ and 9.7 × 10^-10^ for Hi-C and gLink scores, respectively). As an example, we show results for *SFMBT1*, a gene encoding for a histone-binding co-repressor protein, which has previously been associated with BD^58^ (**Fig. 4a**).

**Figure 4.**
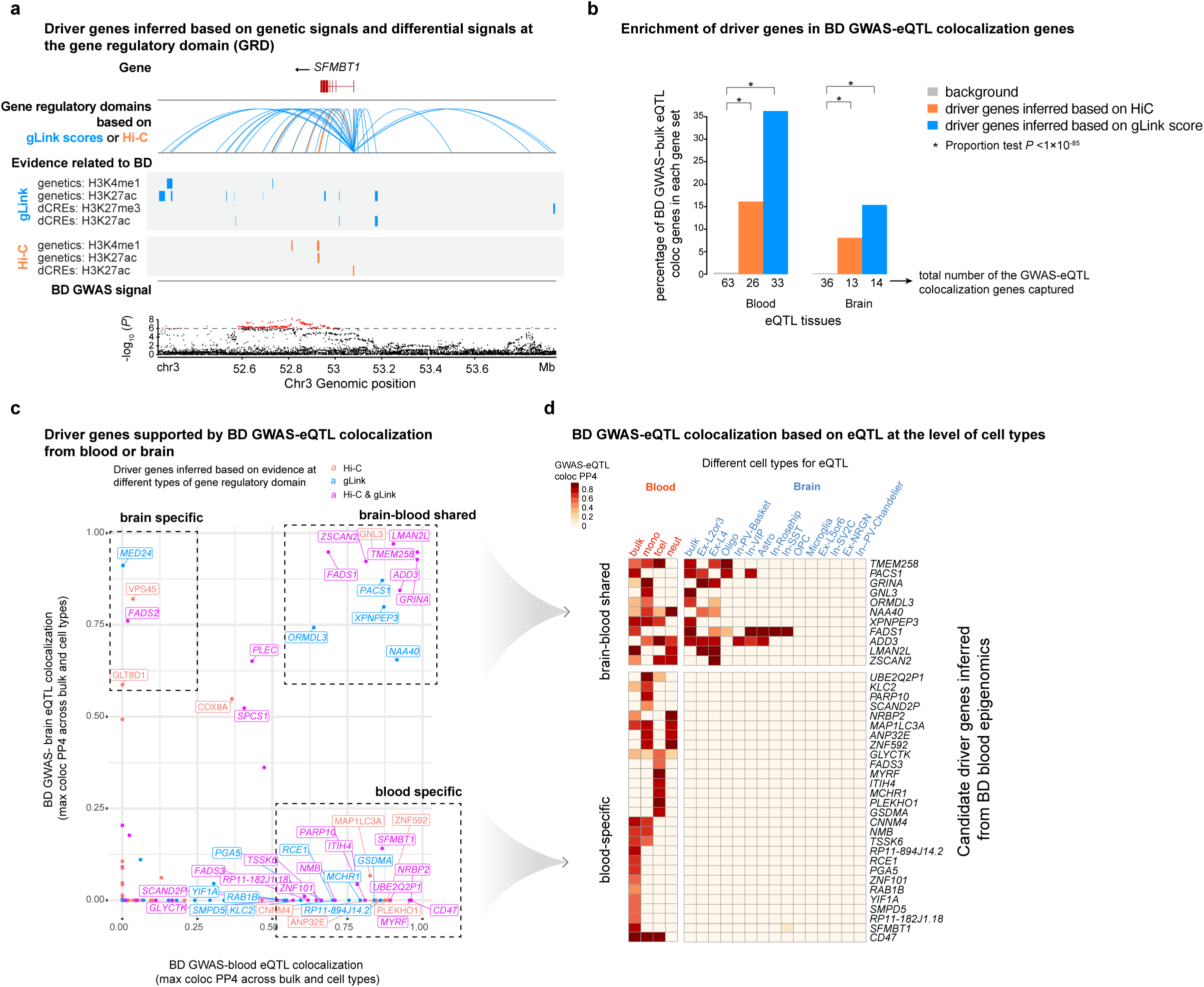
BD Driver genes shared between the blood and brain. **a,** Driver genes inferred based on genetic signals and differential signals at gene regulatory domains (GRD). *SFMBT1*, which was identified as a driver gene in blood for BD based on its links (based on either gLink or Hi-C) to both CREs genetically associated with BD (e.g. genetics: H3K4me1) and BD dCREs (e.g. Dn_H3K27ac), shown as an example. The BD GWAS signal is shown at the bottom panel. **b,** Enrichment of driver genes in BD GWAS–eQTL colocalization genes. The bar plot compares the percentage of BD GWAS–eQTL colocalized genes from the brain or blood in three different gene sets: all eGenes as background, driver genes based on Hi-C linking (in orange), and driver genes based on gLink scores (in blue). Asterisks indicate the significance of the tests comparing different proportions (one-sided proportion test). **c,** Driver genes supported by BD GWAS–eQTL colocalization from blood or brain. The scatter plot shows the BD GWAS–eQTL colocalization signal (max coloc PP4) across brain tissue/cell types (y-axis) and blood tissue/cell types (x-axis), with tissue-specific and shared driver genes highlighted and genes supported by different types of linking evidence in different colors. **d,** BD GWAS–eQTL colocalization based on eQTL at the level of cell types. The heatmap shows the BD GWAS–eQTL colocalization signal (coloc PP4) for driver genes (by row) with strong GWAS–eQTL colocalization signal in the blood (coloc PP4 ≥0.5) across different cell types or bulk tissues (by column). Genes were grouped based on whether there were also strong colocalization signals in any of the brain bulk tissue or cell types.

We next tested whether eQTL signals of BD driver genes from either the blood or brain support their association with BD genetic signals. The driver genes identified via both types of linking evidence showed significant enrichment (one-sided proportion test *P*< 1×10^-85^) for the genes associated with BD genetics based on GWAS–eQTL colocalization in blood or brain bulk samples (*coloc* PP4 ≥0.5) (**Fig. 4b**). The higher enrichment in driver genes based on gLink scores vs. Hi-C data in both tissues also confirmed that the CRE–gene links identified via gLink scores could prioritize genes with stronger disease-associated genetic signals^56^. To increase the power of GWAS–eQTL colocalization, we also included eQTL data at the cell type level for blood from the BLUEPRINT project^59^ and for the brain from a ROSMAP cohort^60^ and carried out the colocalization analysis for the driver genes identified above. We identified 46 driver genes showing GWAS–eQTL colocalization signals in either the blood or brain (*coloc* PP4≥0.5), including 28 genes with colocalization only in the blood (blood *coloc* PP4≥0.5, brain *coloc* PP4<0.25),4 with colocalization only in the brain (brain *coloc* PP4 ≥0.5, blood *coloc* PP4<0.25), and 11 genes with colocalization in both tissues (**Fig. 4c, Supplementary Table 5**). These results indicate that the majority of BD genetic signals captured in our blood samples based on GWAS–eQTL colocalization may specifically function through the circulating immune system rather than in the brain.

We next focused on 39 driver genes supported by GWAS–eQTL colocalization in the blood (*coloc* PP4 ≥ 0.5) and examined them further at the cell type level (**Fig. 4d, Supplementary Table 5**). While the majority of genes were specifically supported by blood eQTL signals, colocalization at the cell-type level also revealed a strong pattern of nine driver genes shared between immune cells and excitatory pyramidal cells of neocortical layers 2/3 (Ex-L2or3) or layer 4 (Ex-L4) in the brain, instead of microglia, the primary brain immune cell type. Interestingly, these neuronal cell types may be causal in BD since they have previously been identified as involved in mediating stress-induced depressive behaviors^61,62^. It indicates these nine genes captured by cell-type-specific eQTL signals from both neuronal and immune cell types may drive BD pathogenesis primarily in neuronal cell types while mirroring genetic effects in immune cells. Our analysis also uncovered genes such as *ORMDL3* and *MAP1LC3A*, which are related to ER stress or calcium signaling (**Fig. 4d**). These pathways are enriched around CREs that potentially share genetic drivers with BD (**Fig. 3d**), and have also been linked to BD^51,63,64^. Furthermore, among the TFs enriched for dCREs (**Fig. 2b**), ATF1, CEBPB, NFIA, and NRF1 are also related to ER stress^65–68^. Together, these results suggest that BD genetic signals may increase the risk of dysregulated ER homeostasis and calcium signaling in the circulating immune system, potentially contributing to BD pathogenesis^12^.

### Heterogeneity of CRE activities in BD patients reveals new phenotypic subtypes

Due to the various risk factors for BD, such as genetics, early exposure to trauma or infection, and comorbidity patterns^69^, BD pathogenesis is believed to be highly heterogeneous^4,6^. To determine whether epigenomic profiles in the blood could reflect such heterogeneity, we first filtered dCREs from each histone mark for robust signals based on dCREs enrichment analysis in chromatin interaction blocks as described above (**Methods**). We then carried out latent factor analysis by MOFA^70^ to capture the variation of the selected dCREs in our cohort (**Extended Data Fig. 5a**) and identified 24 latent factors (LFs) across different histone marks (**Extended Data Fig. 5b**). Next, we investigated whether these latent factors, representing major epigenomic variation across individuals, were driven by any physiological or medical information from the EHR or with the estimated cell fraction variation across all BD patients. Notably, LF2-5 showed a strong correlation with the estimated cell fractions; LF7 was significantly correlated with terms related to infections of a specific organ; LF10 with inflammation, infection, diabetic, and cardiometabolic status-related terms; LF11 with NK cell fractions and cytomegalovirus (CMV) infection, which has been reported to be associated with BD^71^; and LF20 was correlated with digestive system-related medicine records. These correlations suggest that LFs reflect various sources of epigenomic variation across BD patients (**Extended Data Fig. 5c**).

We then clustered all patients and controls with these LFs based on the correlation matrix among them (**Fig. 5a**). There were five groups of individuals showing significantly different proportions of the patients (Chi-squared test *P*=9.1×10^-6^). Group 1 and 2 show higher patient proportions while group 5 shows lower proportion than that of the entire cohort (two-sided proportion test), indicating different BD risks or comorbidity associated with the different groups (**Fig. 5b**). We next investigated whether patients from each group, namely patient subgroups, reflect heterogeneity related to BD pathology or physiology. We annotated each patient subgroup with EHR data to identify differentially enriched terms vs. other groups (**Fig. 5c**). Patients in subgroups 1 and 2 were both enriched for infection- and inflammation-related terms, consistent with the enriched terms for dCREs in BD (**Fig. 2a**). Subgroup 3 showed enrichment of the terms *polyethylene glycol 3350*, which indicates osmotic laxative usage, and *impaired glucose tolerance test (oral)*. Both terms relate to BD: Osmotic laxatives are commonly used to treat constipation, which is one of the major side effects of antipsychotic medicines^72^ and diabetes has been reported to be associated with BD^73^. The prevalence of BD is also significantly higher in irritable bowel syndrome patients, who may present with constipation, than controls^74^. Subgroup 4 showed enrichment of the term *quetiapine_current*, an atypical antipsychotic medicine for BD, and *hypertension*, which BD patients have a higher risk for^75^. We did not identify strong BD-relevant terms for subgroup 5, which also showed the lowest fraction of BD patients compared to other subgroups (**Fig. 5b**).

**Figure 5.**
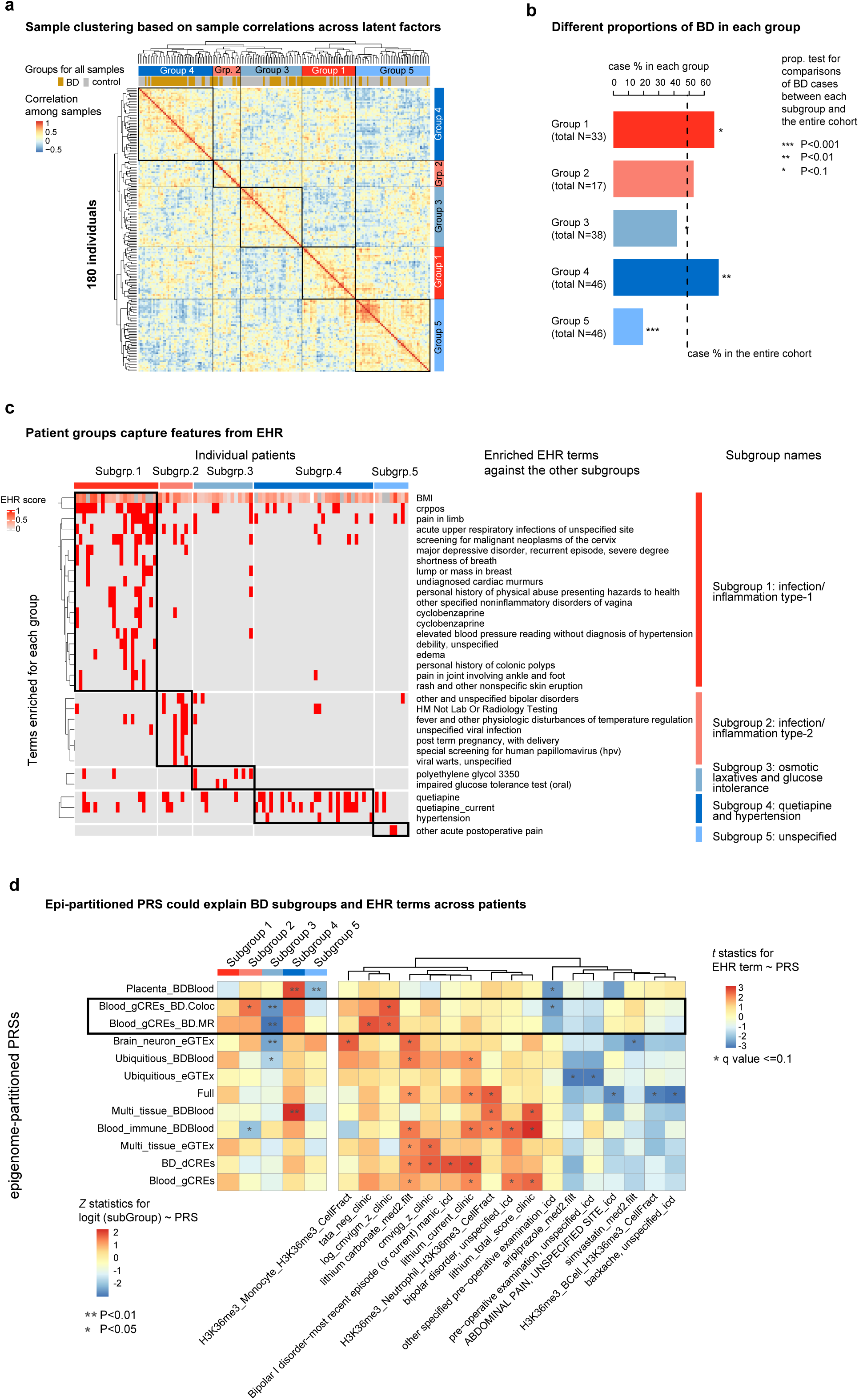
Subtyping of BD patients and annotation of subgroups. **a,** Sample clustering based on correlation across latent factors. The heatmap shows the correlations (Pearson’s correlation coefficient) among patients and controls (by row and by column) based on the latent factors and hierarchical clustering based on the correlation matrix. **b,** Different proportions of BD in each group. Order the same as in panel a. Significance levels of the proportion test to compare BD fractions between group 5 and other groups. **c,** Patient groups capture features from EHR. Top: the heatmap shows the value of latent factors (by row) across individuals (by column) for each patient group; bottom: the heatmap shows the normalized EHR score (1 for presence, 0 for absence for categorical terms, continuous terms are normalized to the range of 0-1) for each individual (by column) for each term (by row) with the group name on the right. **d,** Epi-partitioned PRSs could explain BD subtypes and EHR terms across BD patients. The left heatmap shows the *Z* statistics with binomial regression models that predict the BD subtypes (binary, by column) across individuals based on various epi-partitioned PRSs (by row) correcting for sex, age, and appearance of EHR terms related to infection/inflammation, glucose, hypertension, pain, and abuse. Asterisks indicate the significance levels of the tests. The right heatmap shows the *t* statistics of linear models that predict known factors (by row) across individuals based on various epi-partitioned PRSs (by column) correcting for sex and age. Asterisks indicate adjusted *P*<0.1 based on BH multiple test correction.

To confirm that different patient subgroups show distinct blood epigenomic signatures, we also identified CRE biomarkers across five histone marks for each patient subgroup. The GO annotations of the 12-2200 CRE biomarkers identified for each subgroup revealed biological processes consistent with the EHR terms characterizing individual subgroups (**Extended Data Fig. 5d**). For example, the biomarker CREs (H3K36me3 and H3K4me1) for subgroups 1 and 2 showed enrichment for infection- and inflammation-related terms; the biomarker CREs (H3K27ac) for subgroup 3 showed enrichment for inositol biosynthetic processes and response to lipopolysaccharide, which are both related to diabetes^76,77^; and the biomarker CREs (H3K27ac) for subgroup 4 showed enrichment for cellular responses to thyroid hormone stimulus, which is related to hypertension^78^.

We next explored the genetic basis of these patient subgroups by analyzing the contribution of aggregated genetic signals, specifically, the BD polygenic risk score (PRS; referred to as the “full” model in **Fig. 5d** and **Extended Data Fig. 5e and f**). To facilitate capturing the genetic heterogeneity of this complex disease in finer detail, we also considered subsets of GWAS single-nucleotide polymorphisms (SNPs) based on epigenomic annotations in the PRS models in a manner similar to the computation of pathway-based PRSs^79^. Specifically, we partitioned the PRS by blood CRE groups primarily active in different tissues or cell types (**Fig. 1b**), dCREs (**Fig. 2b**), gCREs (**Fig. 3a**), and CRE sets detected in brain, muscle, heart, and lung samples from a previous eGTEx report, generating “epi-partitioned PRSs” (**Methods**). We found PRSs partitioned by CRE groups primarily active in the immune system, active across multiple tissues, and gCREs identified in our blood study showed significant differences between BD and control individuals (**Extended Data Fig. 5e**). Another two PRSs, which were built based on gCREs associated with BD genetics either by GWAS-QTL colocalization (*blood_gAREs_BD.coloc*) or Mendelian randomization (*blood_gAREs_BD.MR*), are particularly interesting. Though they did not show significant difference between patients and controls, however, these two scores could explain inter-individual variation among BD patients that the full model missed (**Fig. 5d**). For example, these two PRSs distinguished subgroups 2 and 3 from the rest of the patients. Both of them were correlated with the reactivation of cytomegalovirus (term “CMV IgM”) but not with CMV infection (term “CMV IgG”), indicating a genetic component underlying the previously reported association between CMV reactivation and BD^71^. They were also correlated with the epigenomic latent factor LF9 across all patients (**Extended Data Fig. 5f**). Importantly, these pieces of evidence support that these two PRSs, reflecting the BD genetic signals associated with dysfunction in circulating immune cells, could capture epigenomic and phenotypic variations among patients. All these results suggest that the integration of profiles based on five epigenomic marks could capture the heterogeneity of the patients—which may arise from a complex interplay of genetic factors, medicine taken, and physiological conditions—providing a unique perspective for therapeutic intervention.

### BD drug response and repurposing

To reveal immune response to BD interventions, we then identified tens to thousands of CREs for each histone mark affected by medications in BD patients. GO enrichment analysis showed six of these drugs associated with immune-related pathways (**Fig. 6a**). For example, the antipsychotic drug aripiprazole, a dopamine system stabilizer^80^, was strongly associated with innate immune response pathways and inflammation and, to a lesser extent, with adaptive immune response pathways. The anti-seizure drugs topiramate and gabapentin shared with aripiprazole their association with the innate immune response- and inflammation-related pathways, respectively. Quetiapine and trazodone were associated with adaptive immune responses, while divalproex was related to the chemotaxis of leukocytes. These results raise the possibility that these drugs, though designed to target neuronal signaling in the brain, may also affect immune functions either as part of their therapeutic effects or as side effects. Besides immune-related pathways, we also noticed pathways known to be affected by certain BD drugs, such as potassium ion transport associated with quetiapine^81^, and telomere and DNA repair with lithium^82,83^ (**Supplementary Table 6**). These results suggest that the peripheral immune system could provide insights into the physiological responses to, and potential mechanisms of action of, drugs primarily designed for central nervous system diseases.

**Figure 6.**
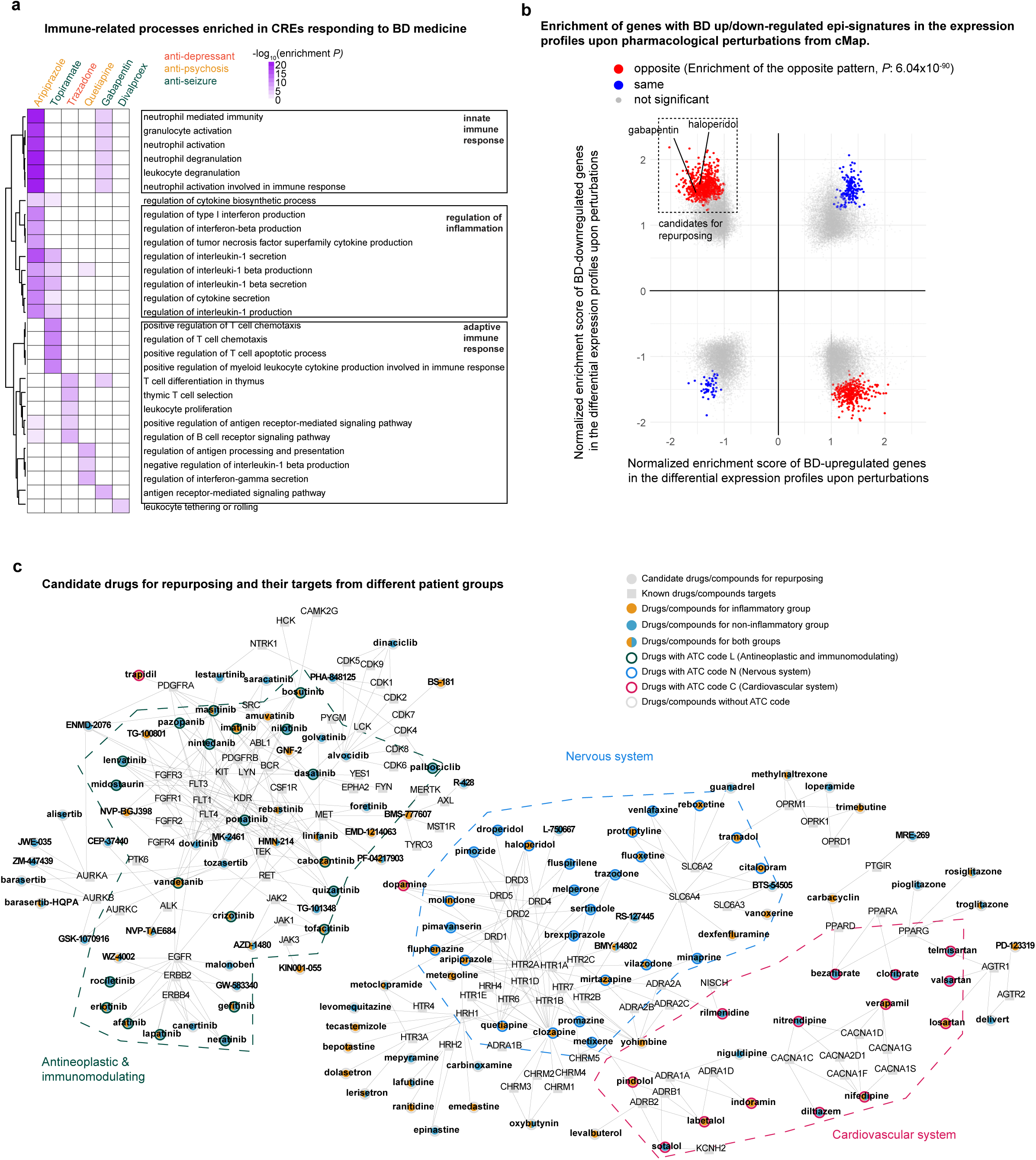
BD drug response and drugs repurposing. **a**, Immune-related processes enriched in CREs responding to BD medicine. The heatmap shows the enrichment (-log_10_ *P*) of biological processes (by row) for each CRE set responding to BD drugs (by column), with different groups of processes highlighted on the left. **b,** Enrichment of genes with BD up/down-regulated epi-signatures in the expression profiles of hematopoietic cell lines exposed to pharmacological perturbations from CMap. Each dot represents an expression profile upon a perturbation; the x-axis and the y-axis represent normalized enrichment scores (NES) for the genes with BD up- and down-regulated epi-signatures, respectively. Colored dots indicate significant enrichment (nominal P<0.001), with red dots representing differential expression profiles with *opposite* directions of NES for BD up- and down-regulated genes (significantly enriched, Fisher’s exact test *P*=6.04×10^-90^), while blue dots representing differential expression profiles with the same directions of NES for the BD up- and down-regulated genes. **c**, Candidate drugs for repurposing and their targets. Nodes represent either drugs (square) or targets (circle) with labels and edges connecting drugs to their targets. The network includes representative connected subgraphs, targeting common mechanisms between inflammatory and non-inflammatory BD subgroups, where at least one drug (in orange) is shared. These subgraphs show a high proportion of drugs originally designed for cancers, nervous system and cardiovascular disorders as labeled.

We then carried out drug repurposing analysis to seek drugs and compounds that could reverse BD-associated immune dysfunctions as represented by the gene sets with up- or down-regulated epi-signatures defined above (**Fig. 2c, Supplementary Table 3**). We applied gene set enrichment analysis (GSEA)^84,85^ to assess the representation of these BD signature genes in the differential gene expression profiles of hematopoietic cell lines treated with 33,627 compounds in Connectivity Map (CMap)^86^ (**Methods**). Out of 49,488 differential expression profiles, 1232 showed significant normalized enrichment scores (NES, nominal *P* < 0.001) for both up- or down-regulated BD signature genes (colored spots in **Fig. 6b**). Further interrogation revealed that a significant majority of these profiles (1014 representing 82% of the total, Fisher’s exact test *P*=6×10^-90^) showed opposite patterns of BD gene enrichment: positive NES for BD-downregulated and negative NES for BD-upregulated genes (red spots in the upper left quadrant in **Fig. 6b**) or vice versa (red spots in the lower right quadrant in **Fig. 6b**), confirming an overall antagonistic relationship between up- and down-regulated BD genes across perturbation conditions. Importantly, 697 of these differential expression profiles, corresponding to 428 compounds targeting 279 proteins, showed an inverse pharmacological impact on BD signature genes, i.e., upregulation of the BD-downregulated and downregulation of the BD-upregulated genes (**purple box in Fig. 6b, Extended Data Fig. 6a, Supplementary Table 5**). These drugs/compounds are particularly noteworthy, since they represent potential candidate agents that could ameliorate BD-associated immune dysregulation.

Next, we tested whether the predicted responsiveness of BD patients to immune cell-targeting drugs and compounds could be further refined by considering BD patient groups established above. We split patients into two groups, an inflammatory group (patient subgroups 1 and 2) and a non-inflammatory group (patient subgroups 3, 4 and 5) (**Fig. 5c**), and identified differential CREs across all five histone marks for each patient group compared to all the controls. We followed the same procedure to identify the genes with BD epi-signatures and then candidate drugs that reversed these BD-associated changes. Similarly to the results from the entire BD patient cohort in **Fig. 6b**, BD genes with up- and down-regulated epi-signatures from both the inflammatory and non-inflammatory groups showed a similarly antagonistic pattern across CMap differential expression profiles (**Extended Data Fig 6. b-c**). We focused on drugs/compounds that reversed patient group-specific BD signature genes in the differential expression profiles (**purple boxes in Extended Data Fig 6. b-c)** and linked them to their known targets in bipartite networks (**Fig. 6c, Extended Data Fig 6. d-e**). Most drugs/compounds and target proteins were either shared by the two groups or connected to the shared ones in various subnetworks, indicating largely shared mechanisms between the two patient groups. We identified a subnetwork (*Nervous system* in **Fig. 6c**) with traditional BD drugs (e.g., aripiprazole) and BD drug targets (e.g., SLC6A4^87^), suggesting that the implicated drugs could reverse BD-associated immune dysfunction in immune cells as a potential mechanism of action. This subnetwork also contained a wide range of receptors and transporters for serotonergic, histaminergic, catecholaminergic, opioid, and cholinergic neurotransmission/signaling. Another subnetwork (*Antineoplastic and immunomodulating* in **Fig. 6c**) with multiple drugs primarily designed for cancers, including various inhibitors of tyrosine kinases FLT3, Aurora Kinases, JAK2, and EGFR, could also modulate immune dysfunctions as expected. We also noticed cardiovascular drugs such as nifedipine, bezafibrate, verapamil, and pindolol, either connected to the previously mentioned *Nervous system* subnetwork or from an independent subnetwork (Cardiovascular system in **Fig. 6c**), which have also been reported to show potential therapeutic effects in BD or related disorders ^88–91^. In each group we also found unique drug–target pairs (**Extended Data Fig 6. d-e**). The list of drugs specific for the non-inflammatory group showed a higher proportion of cardiovascular drugs relative to the inflammatory group (Anatomical Therapeutic Chemical code C, 7 of 24, i.e., 29% vs. 2 of 15, i.e., 13%) (**Supplementary Table 6**). Notably, diosmin, a cardiovascular drug that ameliorated depression behavior in an LPS-induced mouse model^92^, was unique to the non-inflammatory BD patients in our study. Together, these results further highlight the value of leveraging blood epigenomic marks of BD patients for facilitating mechanism-based patient classification.

## Discussion

By analyzing samples and information readily available from living patients (buffy coat cells, genotypes, and EHR data), our study provides significant new insights into the pathomechanisms of BD and the patient heterogeneity in our cohort, thereby also offering a roadmap for similar investigations into other psychiatric disorders. We integrated multimodal data, including 833 ChIP-seq profiles across five major histone marks and genotypes from WGS data, as well as ICD-9 codes, procedure codes, patient-provided information, lab tests, and prescription codes from the EHR of 180 Mayo Biobank participants. We included BD patients and control participants matched to case subjects based on age and gender and lacking any psychiatric condition or disorder. We specifically explored the potential role of circulating immune cells in BD, for which supportive data have been steadily accumulating^13,93,94^. Our analysis identified genes with strong genetic and epigenetic links to BD. Importantly, a large proportion of these genes were linked to BD only by genetic and epigenetic evidence in blood, suggesting that immune function dysregulation in circulating cells may contribute to BD pathogenesis independent of resident immune cells of the brain. We also subtyped patients based on blood epigenomic signals into five subgroups, four of which had strong enrichment of BD-related medical terms. By analyzing the genetic basis of BD patient heterogeneity, we found that BD genetic signals associated with dysfunction in circulating immune cells could capture epigenomic and phenotypic variations among patients. Drug repurposing analysis performed by testing the enrichment of BD epigenetic signature genes in the differential gene expression profiles of hematopoietic cell lines in response to pharmacological perturbation revealed potential peripheral effects of BD drugs and identified drugs/compounds that could ameliorate BD-associated immune dysregulation, including agents with predicted selective effects in patients with or without inflammatory epigenetic profiles. These results highlight that blood epigenomic signals can reflect the physiological state variation of BD patients and are, therefore, promising as biomarkers that could facilitate precision medicine in BD such as matching patients to the right medications.

Immune dysregulation has previously been noted in psychiatric diseases, particularly in bipolar disorder^95^. For example, increased levels of pro-inflammatory cytokines such as IL-1β, IL-6, and TNF-α have been reported in patients with BD^96^ and genetic studies have revealed correlation between BD and autoimmune disorders, such as celiac disease and Crohn’s disease^97^. However, these studies were limited by reliance on a single modality, and the causal relationship between immune dysfunction and BD remained unclear. To address this problem, we tracked the impact of BD-associated genetic variants in immune dysfunction, leveraging multiple data types in the circulating immune system. Most of the BD-associated genetic variants based on GWAS are in the non-coding regions^98^, which makes it challenging to track their actions through their target genes in various cell and tissue types. We applied the following five strategies to address this challenge: (1) Global enrichment: We analyzed genome-wide enrichment of disease-associated genetic variants in CREs primarily active in a specific tissue or cell type^32,99^, suggesting tissues/cell types where these causal variants function (**Fig. 3b**); (2) GWAS–QTL integration: We integrated GWAS signals and molecular QTLs for histone modifications (**Fig. 3c and d)** and gene expression (**Fig. 4c and d**) to specify CREs and genes a given disease-associated genetic variant can exert its effect at the molecular level; (3) Regional regulation: Focusing on chromatin interaction blocks with BD-associated gCREs, we found that these chromosomal units enriched BD dCREs in blood, supporting that immune dysregulation in BD may stem from BD genetic variants (**Extended Data Fig. 4b**); (4) CRE–gene linking: We identified driver genes by linking both BD genetic signals and differential signals through two CRE–gene linking strategies, namely, Hi-C and gLink scores (**Fig. 4a**); and (5) Blood-brain comparison: we compared GWAS–eQTL colocalization for driver genes between blood and brain, decoupling the genes specific to blood from those shared between the two tissues, permitting the identification of BD-relevant immune dysfunctions encoded exclusively in circulating white blood cells (**Fig. 4c and d**). Our systems strategy of integrating information based on genetics and epigenetics provides evidence of the impact of BD-associated genetic signals on the immune system, highlighting candidate SNP–CRE–gene circuits, especially those specific to the circulating immune system, that link immune dysregulation to BD.

Our study provides a comprehensive view of how immune components are involved in BD pathogenesis and affected by BD medication. We discovered upregulated innate immune response and downregulated adaptive immune response pathways in BD, consistent with previous findings of regulatory T cell anergy and senescence in BD^100^. We identified TFs responsible for these alterations, such as FLI1, and their potential connections to BD genetic signals or differential signals in blood. Furthermore, we also identified differentially regulated CREs that may be affected by BD medications, indicating that these drugs may modulate the immune system in distinct ways. We also captured additional upstream CREs inferred from genetic signals. These gCREs, whose hQTL signals were colocalized with BD signals, may indicate a causal effect in circulating immune cells on BD. They are linked based on genetic evidence or chromatin interaction to ER homeostasis and calcium signaling-related genes, including *TMEM258*, *GRINA*, *ORMDL3*, *LMAN2L*, *MAP1LC3A*, *ITIH4*, *MCHR1*, *RAB1B*, *Y1F1A*, and *CD47* (**Fig. 4d**). Importantly, calcium signaling and ER stress were also shown to be connected to the BD downstream events of T cell energy and exhaustion^101,102^. Consistently, we also identified cardiovascular drugs, e.g., nifedipine, targeting calcium signaling pathways, which are predicted to reverse BD associated immune dysregulation (**Figure 6c**). Additionally, we found other potential BD driver genes in the circulating immune system, including genes related to epigenetic regulation (*SFMBT1* and *ANP32E*), apoptosis/DNA damage (*XPNPEP3*, *NRBP2*, *PLEKHO1*, and *PARP10*), autophagosome (*MAP1LC3A*), anti-phagocytosis (*CD47*), and fatty acid metabolism (*FADS1*) (**Supplementary Table 5**).

We next investigated the potential of blood as a surrogate tissue for studying psychiatric disorders. The importance of “liquid biopsies” has been increasingly recognized for their potential for identifying biomarkers in easily accessible tissues. Several studies have been conducted to establish connections between the blood and brain at various molecular levels, such as gene expression, DNA methylation, plasma cell-free DNA methylation, or QTL signals^103–107^. Our findings revealed three potential mechanisms connecting the circulating immune system to psychiatric disorders such as BD: (1) immune dysfunction manifest in BD, such as upregulated innate immunity and downregulated adaptive immunity (**Fig. 2a**); (2) BD-genetics-driven alterations specifically in the immune system, such as genes related to calcium signaling and ER-related transport, potentially exacerbating BD pathogenesis (**Fig. 4d**); and (3) events in the immune system mirroring those in the brain in BD due to shared genetics or epigenetic regulations (**Fig. 4d**). This third mechanism is currently largely under-studied. Specifically, our analysis of cell-type-specific GWAS–eQTL colocalization revealed that a subset of driver genes, some also related to calcium signaling and ER-related transport, were shared between immune cells and specific neuronal cell types: excitatory pyramidal cells of neocortical layers 2/3 (Ex-L2or3) or layer 4 (Ex-L4). These neuronal cell types have been previously implicated in stress-induced depressive behaviors. Interestingly, these genes were not shared with microglia, the primary brain immune cell type. Consistently, ER homeostasis and calcium signaling are also central to neuron homeostasis in the brain and are highly relevant to BD. For instance, studies have shown that disruptions in calcium signaling pathways may contribute to BD pathogenesis by affecting neuronal excitability, plasticity, and survival^108^. These three mechanisms provide various perspectives for identifying biomarkers in blood for psychiatric disorders.

We next used our blood profiles of five histone marks to reveal distinct patient subtypes in our Type I BD cohort, without accessing any clinical information. We grouped all patients using 24 epigenomic LFs and identified five groups by thresholding the hierarchical cluster tree (**Fig. 5a**). Our blood epigenomic subtyping showed significant enrichment of phenotypic, clinical, and treatments features in these patients, highlighting the clinical relevance of our findings (**Fig. 5c**). To distinguish whether these epigenomic LFs were solely a consequence of treatment, or perhaps drivers of disease subtypes leading to treatment differences, we investigated the genetic basis of these patient subgroups. We partitioned the BD PRS by tissue-specific CRE groups or blood gCREs associated with BD genetics and found that the latter (Blood_gAREs_BD.Coloc and Blood_gAREs_BD.MR) could explain inter-individual variation among BD patients including the reactivation of cytomegalovirus (**Fig. 5 d**), revealing a genetic component in the previously reported association between CMV reactivation and BD^71^. These two PRSs also distinguished subgroups 2 and 3 from the rest of the patients. Interestingly, other BD PRSs seem to capture other aspects of patient heterogeneity, for example, whether patients are currently taking lithium or aripiprazole. These novel findings provide additional insights into the potential molecular basis of BD subtypes and their connections to various risk factors and treatments and indicate that genetic signals associated with dysfunction in circulating immune cells can capture epigenomic and phenotypic variations among patients, further supporting a role for immune dysregulation in circulating cells in BD pathogenesis and the utility of analyzing peripheral blood cells to obtain mechanistic insights into BD and to develop biomarkers for patient classification.

We finally aimed to establish a link between peripheral immune cell epigenomics and drug responses in BD. CREs across histone marks in blood in response to patient medications from the EHR revealed specific associations of common BD medications with adaptive and innate immune system pathways and inflammation (**Fig. 6a**), supporting the findings that these drugs, besides targeting central nervous system functions, may also have direct effects on circulating immune cells^109,110^. **Inspired by this finding, we reasoned that drugs or compounds reversing the immune dysregulation may potentially be repurposed for BD.** We thus carried out drug repurposing analysis by testing the enrichment of the blood gene sets with BD up- or down-regulated epi-signatures in the differential expression profiles of hematopoietic cell lines treated by pharmacological agents in CMap. The analysis revealed 428 compounds that repressed genes with BD up-regulated signatures and activated genes with BD down-regulated signatures, suggesting that they could ameliorate BD-associated immune dysregulation (**Fig. 6b**). Further refining these studies by considering patient groups with or without inflammation-related EHR signatures uncovered subnetworks of drugs/compounds and drug targets related to the nervous system including well-established BD drug targets such as serotonin receptors and transport mechanisms (**Fig. 6c**), consistent with the result of BD medicine response analysis mentioned above. The subnetworks also include tyrosine kinases with roles in immune cells and cancers, and cardiovascular drugs including some with potential therapeutic effects in BD. Interestingly, non-inflammatory patient group showed a higher proportion of cardiovascular drugs relative to the inflammatory group (**Supplementary Table 6**). These data suggest that epigenomic analysis in peripheral blood immune cells could identify new interventions and help match BD patients with the right therapies in the context of immune modulation.

Our study has several limitations. First, we could not include gene expression data since the common protocol used for the biobanking of buffy coat samples was designed to preserve DNA, not RNA. However, we overcame this by using epigenomic signals, especially H3K36me3, a mark for the gene body, to infer differential gene expression withby meta-analysis. Second, our analyses were not performed at the single-cell or cell-type level. Nevertheless, we used deconvolution analysis to ensure that cell fraction changes were accounted for during our differential signal detection and QTL mapping. Additionally, we did not have access to paired brain postmortem samples to check the brain–blood sharing of the differential signals. In future studies, a paired-sample design would be more powerful to detect blood biomarkers that recapitulate changes in the brain. We also did not have drug response or other longitudinal data. We plan to integrate these data types in future work to better understand whether our subtyping is stable throughout an individual’s lifespan and meaningful for predicting drug response. Finally, our work only addressed drug effects in the immune system. A more accurate prediction would require integrating the peripheral and central nervous system responses. We also noticed that some of the suggested drugs might lead to detrimental, rather than beneficial effect to BD. For example, flutamide may induce manic-like episodes^111^ (**Extended Data Fig. 6d**) and nicotine could exacerbate manic episodes in individuals with BD^112^ (**Extended Data Fig. 6e**). Conversely, peripheral blood epigenomic profiling may also be useful for linking medications to adverse effects on BD.

Studies like ours are essential for understanding psychiatric diseases because they provide a comprehensive view of the molecular mechanisms and patient heterogeneity. By integrating multimodal data and investigating the immune components of BD from both genetic and epigenetic perspectives, this work offers valuable insights into the potential role of circulating immune cells in BD pathogenesis. Furthermore, our findings on BD subtypes based on blood epigenomic signals pave the way for improved patient monitoring and personalized treatment recommendations. Despite the limitations, our study represents a significant step toward understanding the complex relationships between the blood and brain in psychiatric disorders such as BD.

## Data availability

Supplementary info and datasets are linked from https://data.broadinstitute.org/compbio1/BDBloodEpigenome/data. Since the raw sequencing data with genetic information are protected, application and authentication are needed before accessing the data.

## Code availability

All codes for this study are available here https://data.broadinstitute.org/compbio1/BDBloodEpigenome/code.

## Author Contributions

This study was designed by M.K., T.O., M.A.F., and J.B., and directed by L.H., T.O., and M.K. Experiments were carried out by S.W.L. under the supervision of J.H.L. and T.O.; L.H., Y.L., X.X., Y.T., and Y.P. performed analyses and interpreted results with help from M.K., T.O., M.A.F, and J.B. Key resources were provided by M.A.F., J.B., E.J., and J.E.O. All authors participated in the discussion of the project. L.H. wrote the manuscript with help from Y.T., A.G, T.O., and M.K.

## Acknowledgement

The authors would like to thank all the donors participating in the study. This work was supported by the Mayo Clinic Center for Individualized Medicine. This study was supported, in part, by the Marriott Family Foundation, the Thomas and Elizabeth Grainger Fund in Bipolar Disorder Novel Therapeutics and Advanced Diagnostics, and the Mayo Clinic Center for Individualized Medicine; none had a role in the design, conduct, analysis, or submission of the study, collection, management, analysis, and interpretation of the data; preparation, review, or approval of the manuscript; and decision to submit the manuscript for publication. It was also supported by NIH grants DK126827 and DK84567 (Epigenomics and Spatial Biology Core) to T.O., and AG058002, MH109978, and MH119509 to M.K.

## Extended Data Figure Legends

**Extended Data Figure 1.**
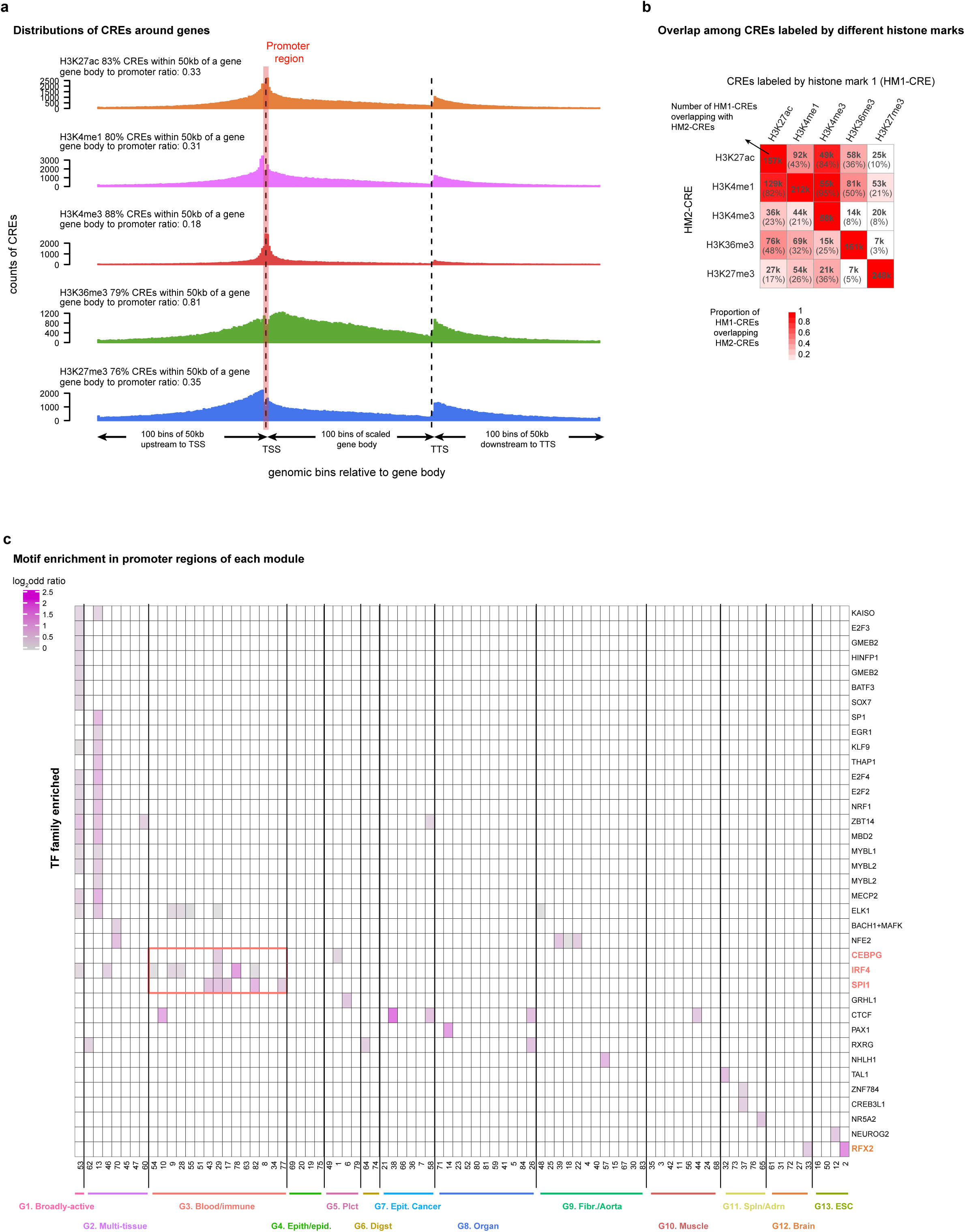
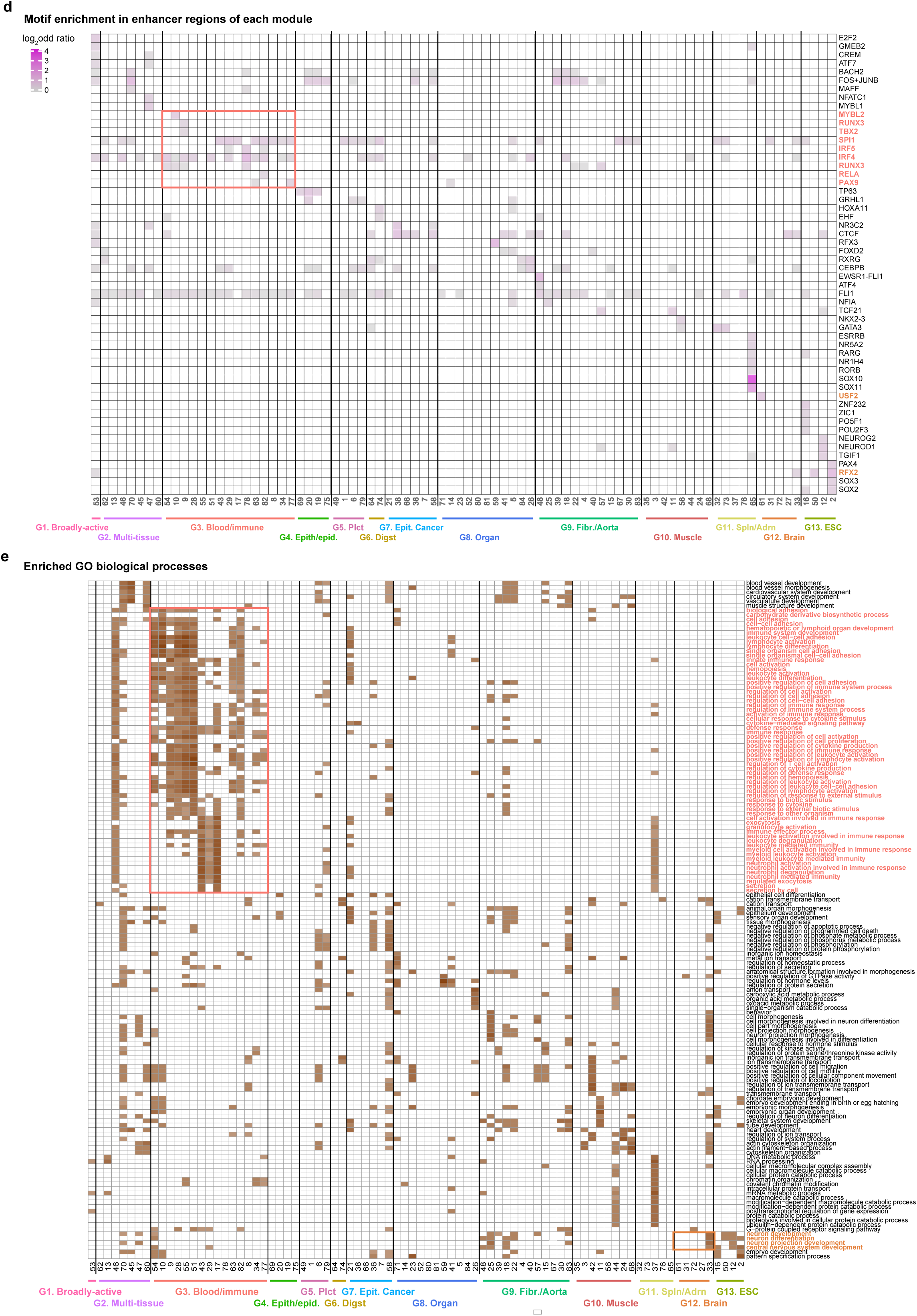
Epigenomic signals and CRE modules. **a,** Distributions of CREs around genes. The bar plot shows the counts of CREs (y-axis) at each genomic bin (x-axis) upstream of the TSS, between the TSS and the TES, or downstream of the TES for different histone marks. **b,** Overlap among CREs labeled by different histone marks. The heatmap shows the proportion of CREs associated with the first histone mark (by column) and the overlapping CREs associated with the second histone mark (by row). Both numbers and proportions of overlapping signals are shown in each cell. **c,** Motif enrichment in promoter regions of each module. The heatmap shows the enrichment (log_2_ odds ratio) of TF motifs (by row) in CREs in promoter regions for each module defined in Fig. 1b (by row), with the TFs enriched in immune modules highlighted. **d,** Motif enrichment in enhancer regions of each module. Same as panel b except for showing CREs related to enhancers. **e,** Enriched GO biological processes. The heatmap shows the enrichment (log_2_ fold enrichment) of GO terms (by row) for each CRE module.

**Extended Data Figure 2.**
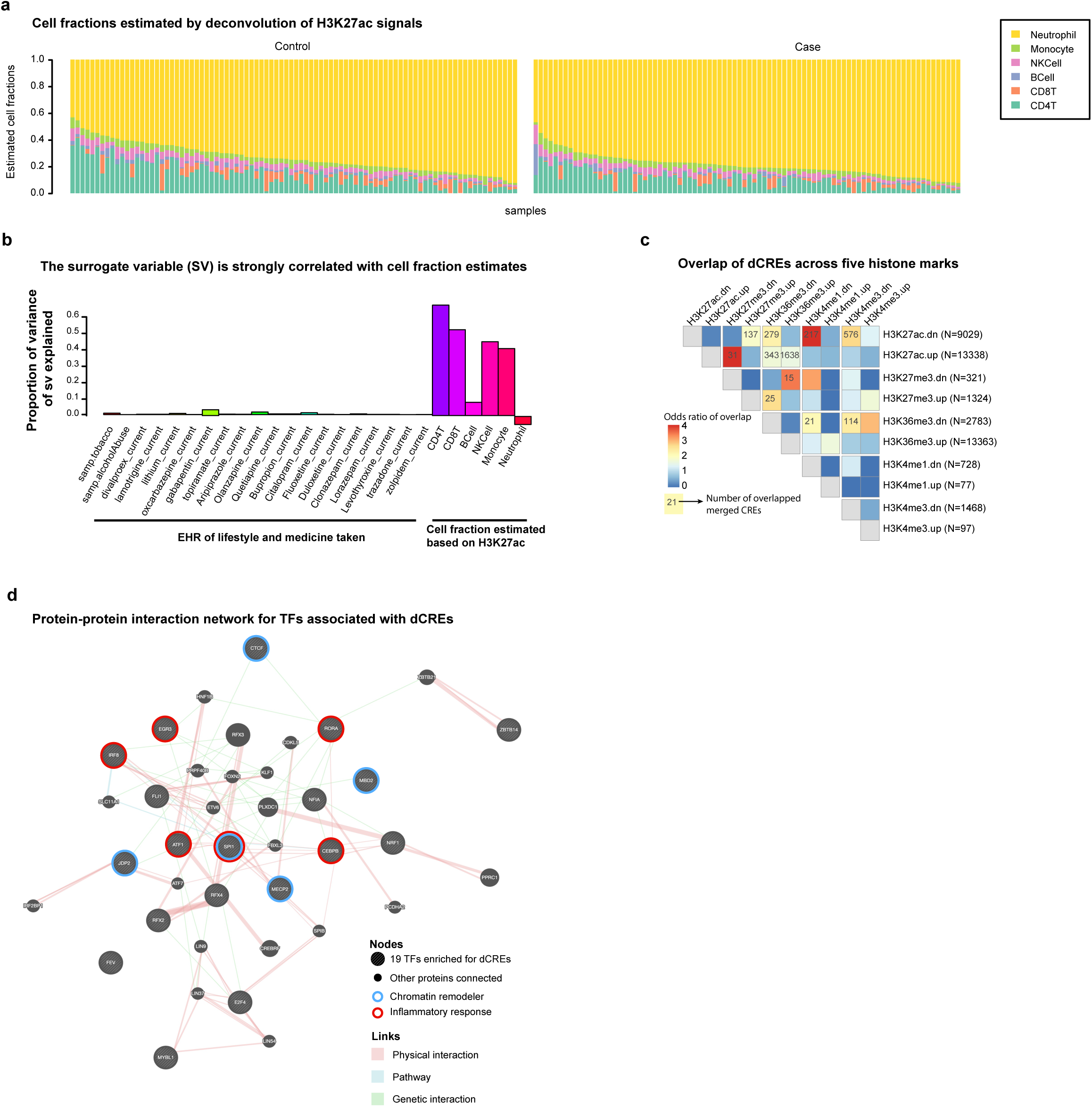
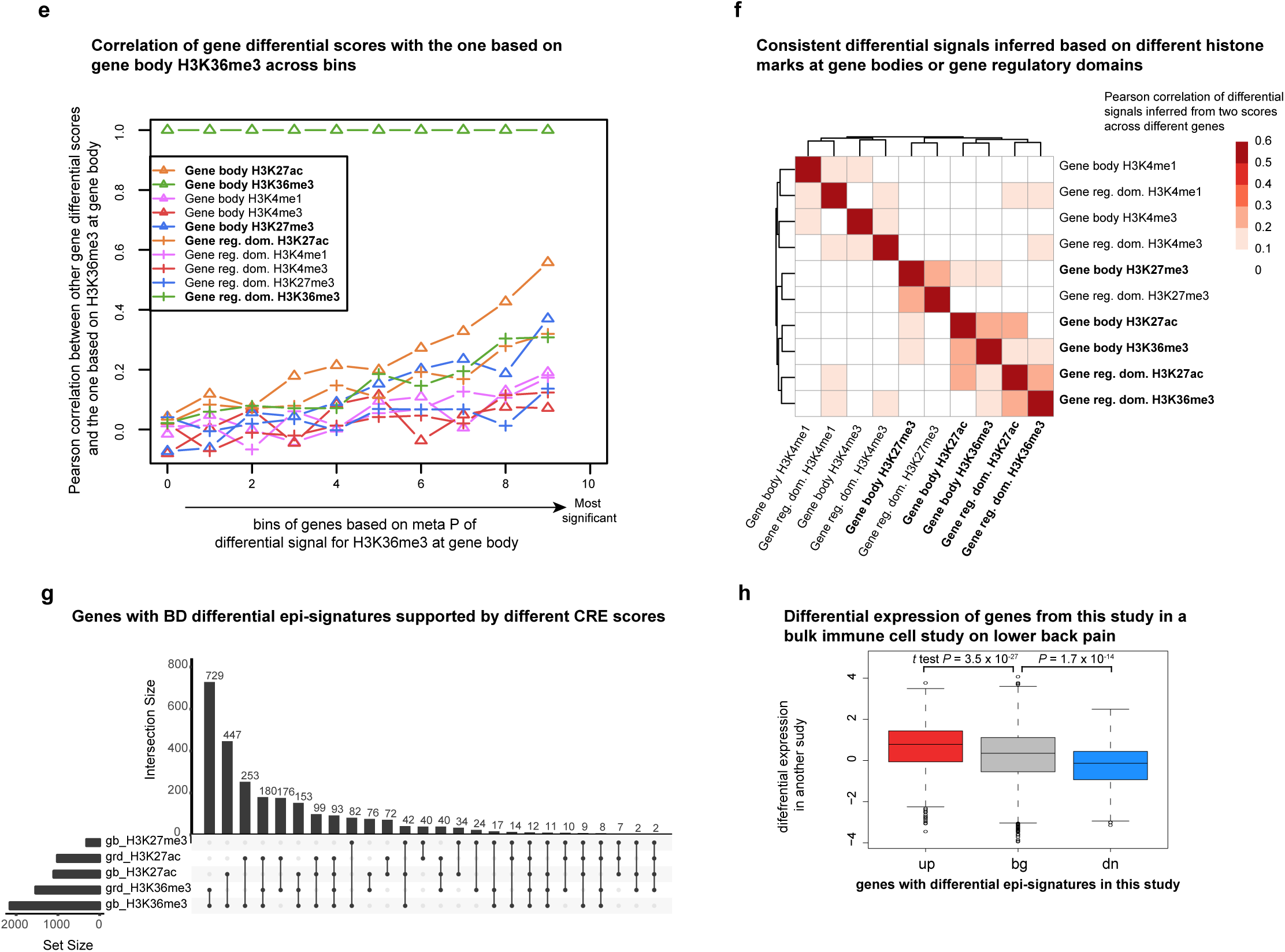
Differential CREs and associated TFs. **a,** Cell fractions estimated by deconvolution of the H3K27ac signal. The bar plots show the estimated cell fraction (y-axis) for controls and patients with different cell types in different colors. **b,** The surrogate variable (SV) is strongly correlated with the cell fraction estimates. The bar plot shows the proportion of variance of SV explained (y-axis) by the different covariates (x-axis). **c,** Overlap of dCREs across five histone marks. The heatmap shows the odds ratio of overlap between each pair of BD dCRE sets with the size of overlapping dCREs shown in the cell if significant (Fisher’s exact test, BH multiple test correction, adjusted *P*<0.05). **d,** Protein-protein interaction network for TFs associated with dCREs. The network shows linking among the TFs associated with BD dCRE sets (big, borders in different colors indicating different functions) and other interacting proteins (small in size). **e,** Correlation of gene differential scores with the one based on gene body H3K36me3 across bins. The scatterplot shows the correlation (y-axis) between gene body H3K36me3 score (z score) with each of the remaining scores for each bin of genes based on the significance of gene body H3K36me3 score (-log_10_ *P* based on a meta-analysis of dCREs). **f,** Consistent differential signals inferred based on different histone marks at gene bodies or gene regulatory domains. The heatmap shows the correlation among scores based on dCRE meta-analysis (by row and by column) included in panel e. **g,** Genes with BD differential epi-signatures supported by different CRE scores. The upset plot shows the counts (y-axis) of genes with BD differential epi-signatures supported by different dCRE meta-analysis scores (x-axis). **h,** Differential expression of genes from this study in a bulk immune cell study on lower back pain. The box plot shows the differential signal in another study (y-axis) for genes identified above with up-regulated epi-signatures (up), genes with down-regulated epi-signatures (dw), and all genes as background (bg).

**Extended Data Figure 3.**
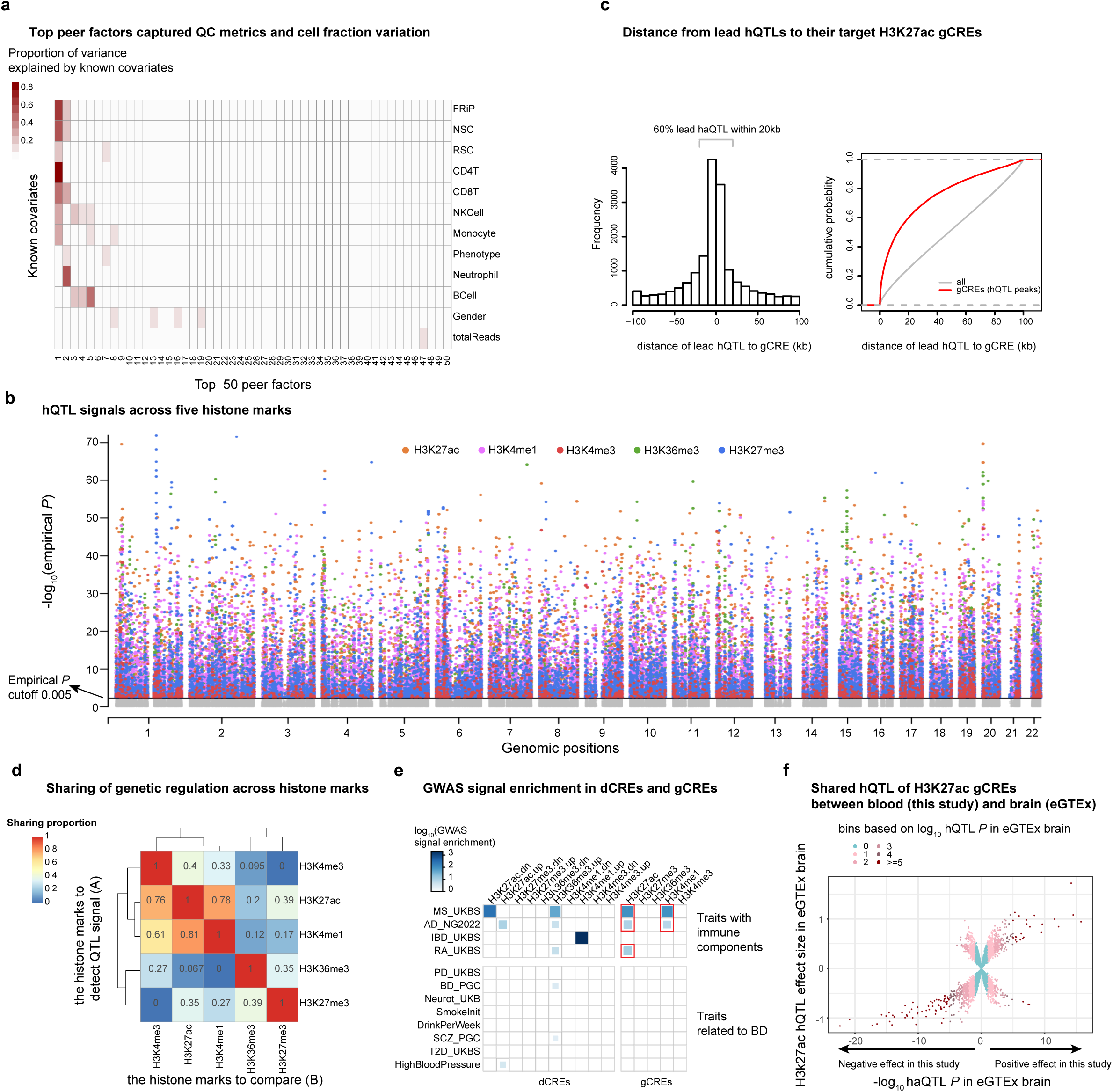
Genetic regulation of CREs and implications in BD genetics. **a,** Top peer factors captured QC metrics and cell fraction variation. The heatmap shows the correlation between peer factors (by column) and known or inferred covariates (by row) for H3K27ac as an example. **b,** hQTL signals across five histone marks. The scatter plot shows the QTL signals (empirical *P*, y-axis) over the genome (x-axis) for five histone marks (in different colors shown on top). **c,** Distance from the lead hQTLs to their target H3K27ac gCREs. Left: the barplot shows the frequency of the top hQTLs (y-axis) across the bins of distance between the QTLs and their target gCRE; right: the plot shows the cumulative distributions of distances between the top hQTLs and its target CRE for gCREs (red) and all CREs as background (gray). **d,** Sharing of genetic regulation across histone marks. The heatmap shows the sharing of hQTLs detected for histone mark A (by row) with those detected for histone mark B (by column). **e,** GWAS signal enrichment in dCREs and gCREs. **f,** Comparing genetic regulation of H3K27ac between blood (this study) and brain (eGTEx). The same plot as panel c, except for showing the comparison of H3K27ac hQTLs between brain and blood. **g,** BD GWAS–hQTL colocalization and hQTL sharing between blood and brain.

**Extended Data Figure 4.**
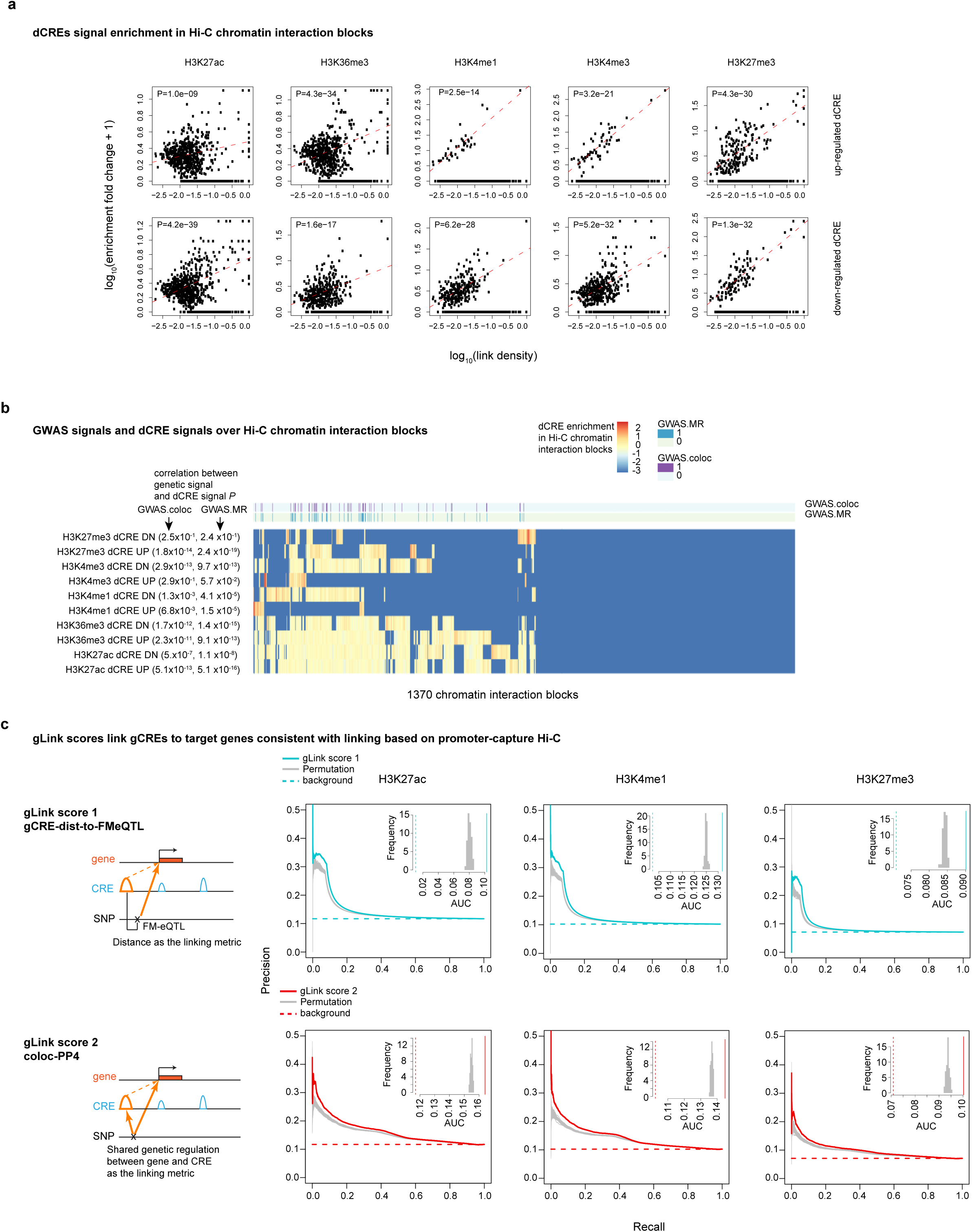
Patterns of dCREs at Hi-C block level and comparison between gLink scores and Hi-C links. **a,** dCRE signal enrichment in Hi-C chromatin interaction blocks. The multi-paneled scatter plot compares the enrichment of dCREs to the density of links—defined as the number of observed Hi-C links divided by the number of all pairs of chromatin regions— across each Hi-C block (each point) for up- or down-regulated dCREs (by row) for each histone mark (by column). **b,** GWAS signals and dCRE signals over Hi-C chromatin interaction blocks. The heatmap shows the enrichment of dCRE sets (by row) in each Hi-C block (by column) with BD genetic signals for each block annotated at the top. **c,** gLink scores link gCREs to target genes consistent with linking based on promoter-capture Hi-C. AUPRC plots show the performance of each gLink score (by row) for each histone mark (by column) with links based on promoter-capture Hi-C data as the benchmark data. Schematic plots to explain two different gLink scores are shown on the left.

**Extended Data Figure 5.**
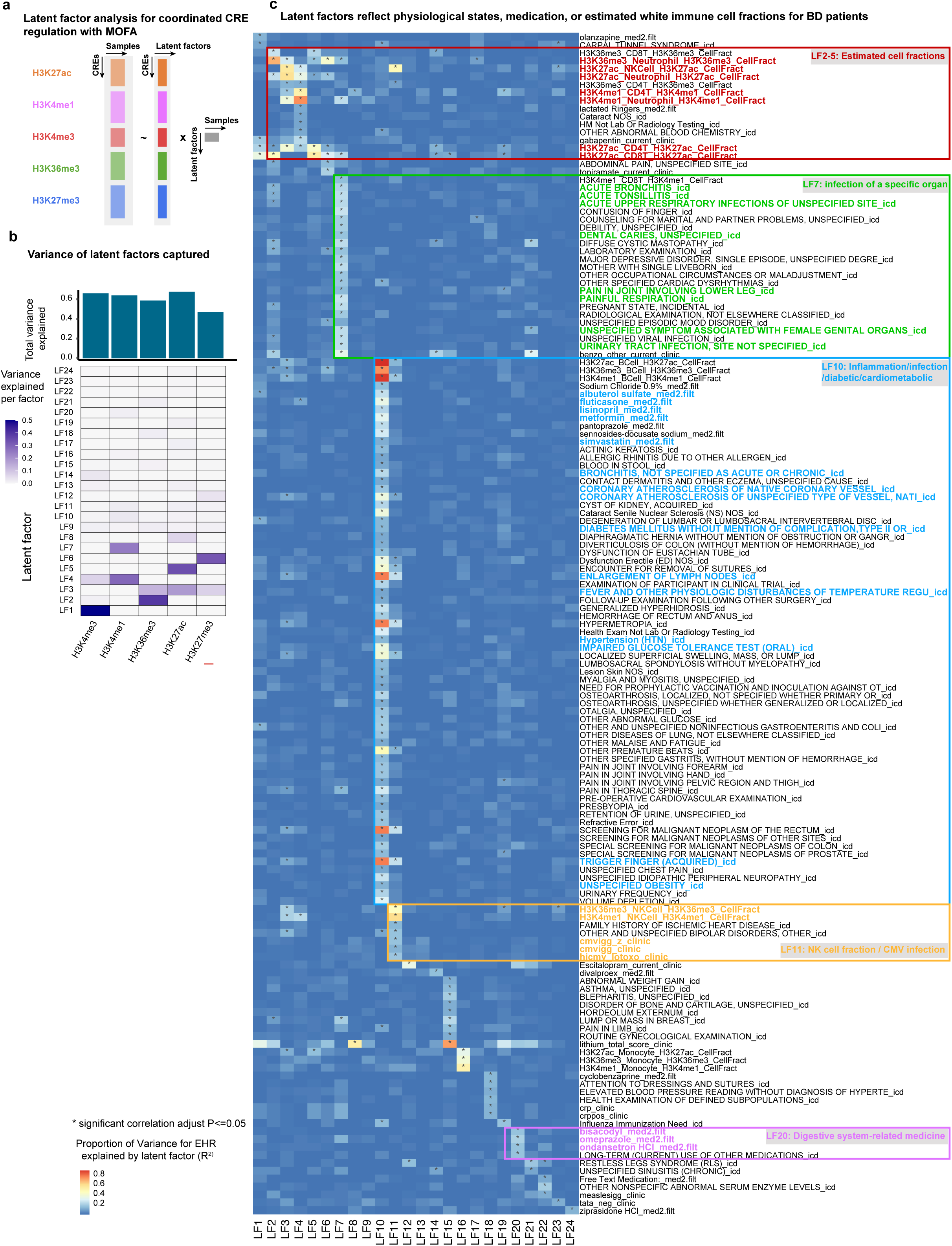
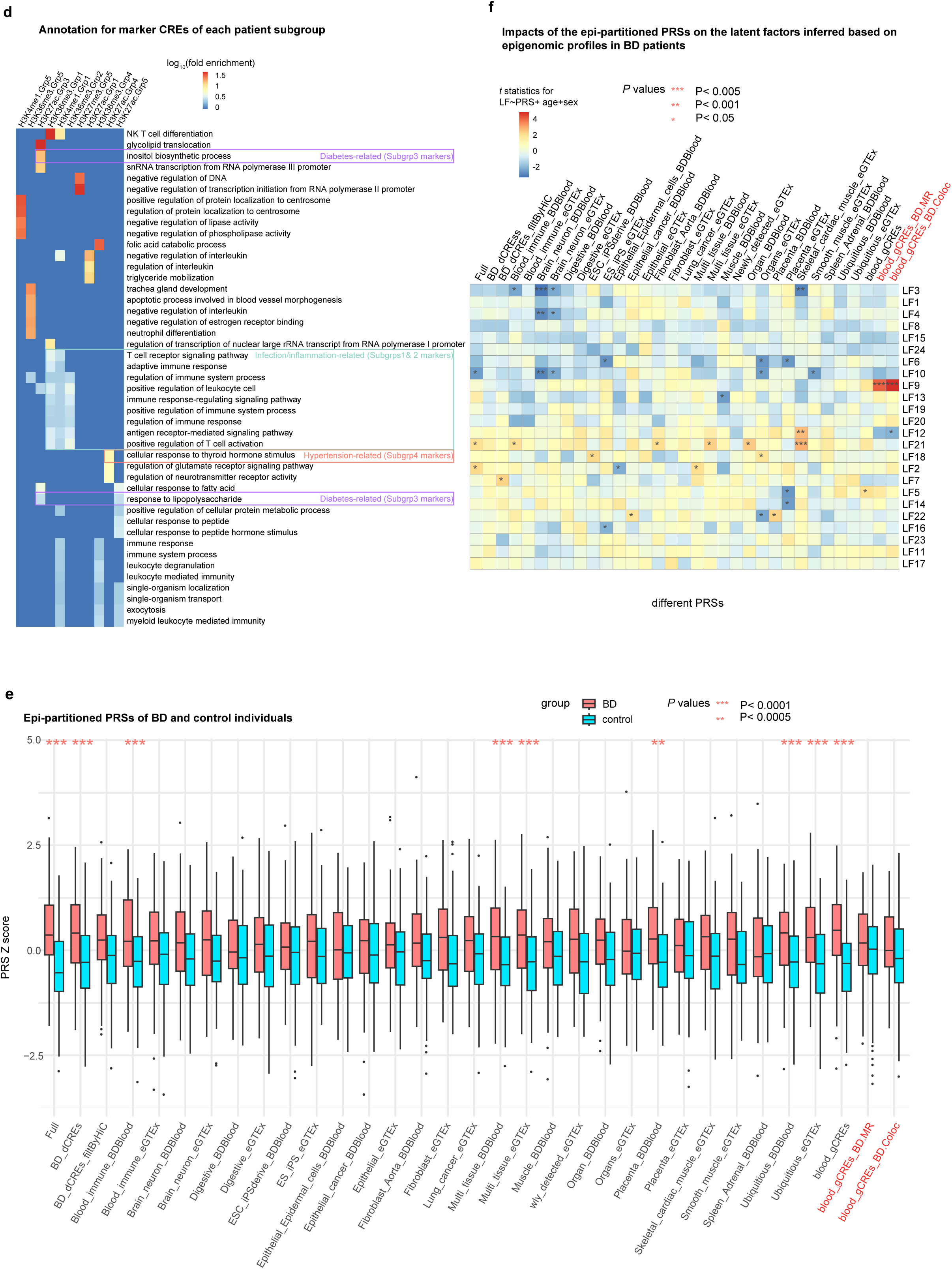
Latent factor analysis and marker CREs for BD subgroups. **a,** Latent factor analysis for coordinated CRE regulation with MOFA. A schematic diagram showing how latent factor analysis based on MOFA works. **b,** Variance of latent factors captured. Top: the bar plot shows the total variance explained by all latent factors (y-axis) for each histone mark (x-axis); bottom: the heatmap shows the variance explained for each factor (by row). **c,** Latent factors reflect physiological states, medication, or estimated white immune cell fractions for BD patients. The heatmap shows the proportion of variance for each covariate/EHR term (by row) explained by each latent factor (by column), with covariates/terms associated with several latent factors highlighted on the right. **d,** Annotation of marker CREs of each patient group. The heatmap shows the enrichment of GO terms (by row) for each marker CRE set identified for each patient group (by column), with terms consistent with prior knowledge highlighted on the right. **e,** Epi-partitioned PRSs of BD and control patients (red and blue box plots, respectively). The x-axis represents different scores; the y-axis shows z scores for each PRS for each individual. Boxes: 25%-75% percentile (i.e. inter-quartile range; IQR); line: median; whiskers: 1.5 IQR. Asterisks indicate the significance levels of the difference of scores between the two groups. **f**, Impacts of the epi-partitioned PRSs on the LFs inferred based on the epigenomic profiles of BD patients. Color of the heatmap represents the *t* statistics of a PRS in a linear regression model to predict a latent factor correcting for age and sex; each row of the heatmap represents a latent factor; and each column represents a PRS, with asterisks indicating the significance levels of the *t* statistics.

**Extended Data Figure 6.**
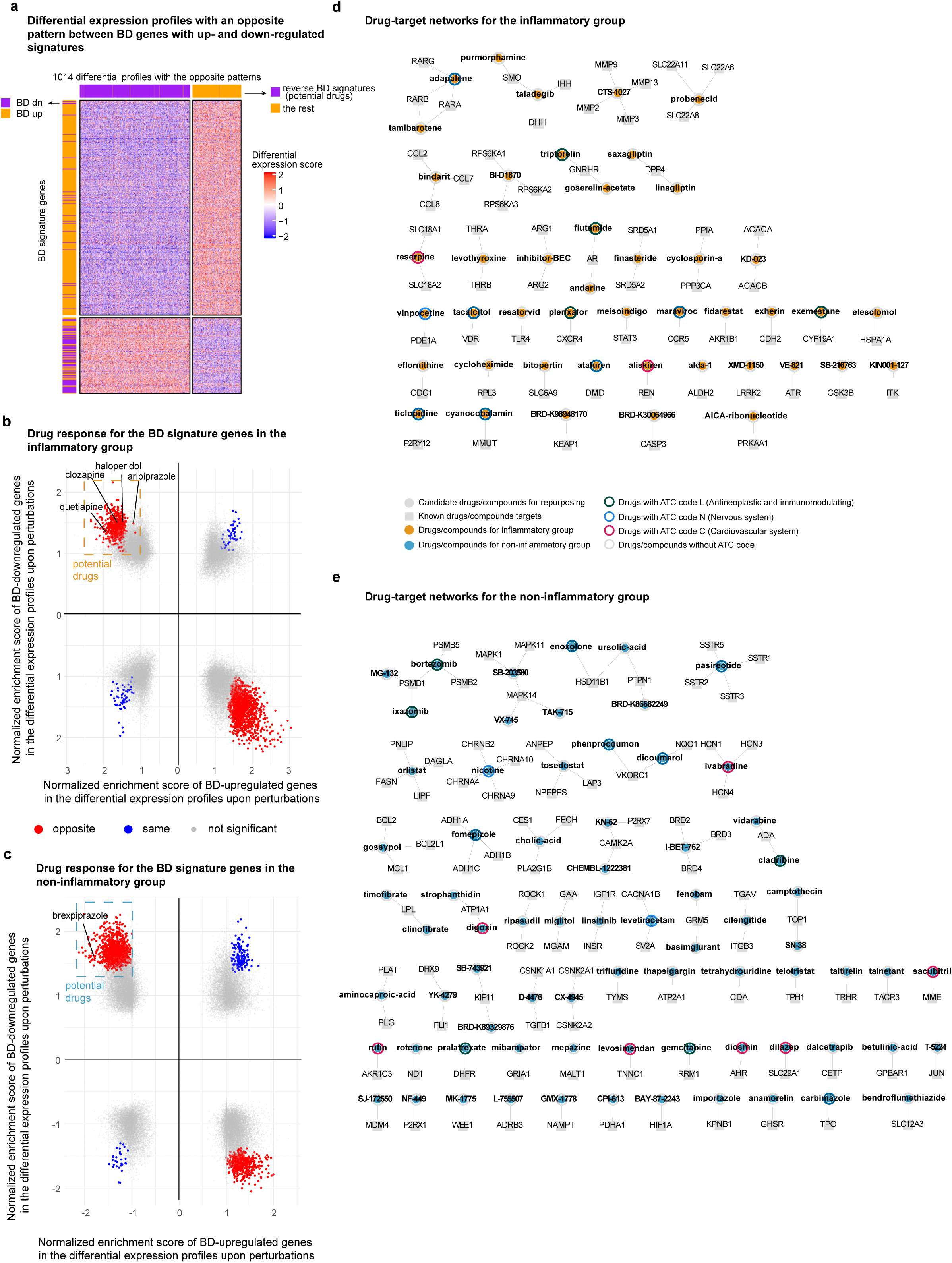
**a,** Differential expression profiles in response to pharmacological interventions (from CMap) with an opposite enrichment pattern for BD genes with up- and down-regulated epigenomic signatures. Rows represent 1014 expression profiles, while columns represent the top 500 differentially expressed genes with BD up-/down-regulated epigenomic signatures. **b**, Enrichment of genes with BD up/down-regulated epi-signatures in the differential expression profiles of hematopoietic cell lines exposed to pharmacological perturbations for the inflammatory subgroups (subgroup 1 and 2 in Figure 5c). BD epi-signatures were identified by comparing the BD patients from inflammatory subgroups to all the controls. Each dot represents an expression profile upon a perturbation; the x-axis and y-axis represent normalized enrichment scores (NES) for the genes with BD up-regulated and down-regulated epi-signatures, respectively. Colored dots show differential expression profiles with significant NES for both gene sets (nominal *P*<0.001), with red dots representing changes with opposite signs of enrichment for BD up- and down-regulated genes and blue dots representing changes with the same signs of enrichment. **c,** The same plot as a except that it is for the non-inflammatory subgroups (subgroup 3-5 in Figure 5c). **d&e,** Network view of repurposed drugs and their targets specifically for inflammatory (d) and non-inflammatory (e) subgroups. Nodes represent either drugs (square) or targets (circle) with labels and edges connecting drugs to their targets.

## Methods

### Mayo Clinic participants

The 180 participants (88 BD cases and 92 age and gender-matched controls) were from the Mayo Clinic Bipolar Disorder Biobank and the Mayo Clinic Biobank^113^, respectively. They had consented to research, including research using their EHR data^26^. This study was reviewed and approved by the Mayo Clinic Institutional Review Board and by the access committees from both biobanks. The EHR information for each of the patients covered five categories, including ICD-9 codes, procedure codes, patient-provided information, lab tests, and medication prescriptions. For control participants, subjects from the Mayo Clinic Biobank were matched on age and gender with BD cases and filtered based on psychiatric conditions in self-reports and/or EHR.

### Common variants from whole-genome sequencing (WGS)

WGS of our cohort (N=179 with one individual missing) were carried out using the same experimental protocol and quality control procedure as in a previous study^114^. In short, DNA from blood samples underwent shearing using a Covaris LE220 ultrasonicator to obtain fragments of about 350bp in size. The fragmented DNA was processed using the NEBNext® DNA Library Prep Master Mix Set tailored for Illumina®. Unique in-house primers were employed to barcode the post-ligated samples, which were subsequently amplified over 6 PCR cycles with the KAPA HiFi HotStart Ready Mix. Library concentrations were gauged using the Qubit® 2.0 Fluorometer. Meanwhile, the quality of these libraries was assessed using a DNA 5K chip in conjunction with a Caliper GX. Precise quantification was ascertained via the qPCR-based KAPA Biosystems Library Quantification kit. Each sample underwent sequencing on a single lane of Illumina’s HiSeq X device using v2 flow cells and reagents to achieve 30X genomic coverage.

The derived fastq files were then channeled through the *Mayo Genome GPS* v4.0 system. Using the Burrows–Wheeler Aligner*^115^*, reads were aligned to the human reference sequence (GRCh38 build). The Genome Analysis Toolkit (GATK)*^116^* was employed for local realignment surrounding indels. Variant identification was executed using the GATK HaplotypeCaller, followed by variant recalibration (VQSR) in line with the GATK’s recommended best practices^117,118^. Genotypic determinations with GQ below 10 and/or depth (DP) lesser than 10 were omitted. Variants with an ED exceeding 4 were excluded from further analyses. We retained only those variants that cleared the VQSR and exhibited a call rate surpassing 95%. Ultimately, our study concentrated on 6,790,411 common variants with a minor allele frequency greater than 5%.

### ChIP-seq experiment

Buffy coat samples isolated from 5-10 ml of peripheral blood were preserved in anti-freeze media (90% fetal bovine serum and 10% dimethyl sulfoxide) at -80 °C. The samples were thawed on ice, cross-linked with formaldehyde (final concentration: 1%) for 10 min at room temperature then quenched with 125 mM glycine for 5 min. Fixed cells were washed twice with 1X TBS and lysed in cell lysis buffer (10 mM Tris-HCl, pH 7.5, 10 mM NaCl, 0.5% IGEPAL) on ice for 10 min. The lysates were washed with 1ml MNase digestion buffer (20 mM Tris-HCl, pH 7.5, 15 mM NaCl, 60 mM KCl, 1 mM CaCl_2_), resuspended with 500 ul of MNase digestion buffer, and incubated for 20 minutes at 37 °C in the presence of MNase (New England Biolabs, Ipswich, MA). After adding the same volume of sonication buffer (100 mM Tris-HCl, pH8.1, 20 mM EDTA, 200 mM NaCl, 2% Triton X-100, 0.2% sodium deoxycholate), the lysate was sonicated for 30 min (30 seconds on, 30 seconds off) in Diagenode bioruptor and centrifuged at 15,000 rpm for 10 min. The supernatant was divided into five microcentrifuge tubes and incubated with ChIP-seq-validated antibodies on a rocker overnight. The following antibodies were used in the experiment; anti-H3K27ac (CST #8173), anti-H3K4me1 (Abcam ab8895; CST #5326), in-house generated anti-H3K4me3 (EDL lot 1), anti-H3K27me3 (CST #9733), and anti-H3K36me3 (Active Motif #61101) (see **Supplementary Table 1**). Immunoprecipitation and library preparation were performed as described^119^. The libraries were sequenced to 51 base pairs from both ends on an Illumina HiSeq 4000 instrument in the Mayo Clinic Center for Individualized Medicine Medical Genomics Facility.

### Quality control of H3K27ac ChIP-seq and preprocessing

We carried out peak calling with the ENCODE pipeline (https://github.com/kundajelab/chipseq_pipeline). Briefly, we mapped reads to human genome assembly hg19 (*bwa*, v0.5.9)^115^, filtered low-quality and multiple-mapped reads by *samtools* (v1.3.1)^120^, and applied *MACS2* (v2.1.1)^121^ for peak calling. Samples were then filtered based on three QC metrics: a) Relative Strand Cross-correlation coefficient (RSC), a key ChIP-seq QC on signal/noise; b) total reads; and c) correlation between the signal of samples and those from blood samples in Roadmap reference epigenome. We define “Tier 1” samples as those with RSC score ≥ 2, total reads ≥ 30M, and for which their matched Roadmap tissue was in the top three most correlated Roadmap epigenomes. We defined "Tier 2" samples with relatively permissive cutoffs: RSC score ≥ 1, total reads ≥ 20M, and the same criteria of matched-tissue correlation.

We used high-quality samples of “Tier 1” to generate a reference peak set for each histone mark. We first selected peaks (q-value ≤0.05) from each sample and then merged them together. Only peaks present in at least two individuals and overlapping with peaks from immune cell samples in Roadmap are kept in consensus peaks sets for each histone mark, resulting in 156,967 (H3K27ac), 247,775 (H3K27me3), 161,248 (H3K36me3), 211,919 (H3K4me1), and 57,811 (H3K4me3) peaks, defined as *cis-*regulatory elements (CREs).

We extracted peak intensity as CRE activity across all "Tier 2" samples for downstream analyses. We defined peak intensity as the number of reads bases within each peak as in a previous report^56^. Peak intensity was then normalized with three steps: 1) to take into consideration library reads depth, first peak intensities are normalized with a factor for each sample to reach 30M total reads across all the peaks identified; 2) peak intensity mean is then defined by peak intensity divided by peak length, and further normalized across samples by dividing the size factor inferred from R package DEseq2^122^, 3) finally peak intensity mean is corrected for GC content using R package *gcapc^123^*.

### CRE modules and functional annotation

We investigated the co-activity pattern of 223,573 CREs annotated by three enhancer/promoter-associated marks (H3K27ac, H3K4me1, and H3K4me3) across reference epigenomes based on H3K27ac intensity as in a previous study^56^. Briefly, we extracted -log_10_P signal from 242 EpiMap H3K27ac samples for each CRE (bigWigAverageOverBed^124^, v2), binarized the matrix by a cutoff of 2, kept CREs with non-zero values in at least one example (N=168,801), and clustered them by a k-centroids algorithm implemented in R package *flexclust* (V1.4-0)^125^ with the Jaccard index as the distance metric and cluster number set as 84 (corresponding to 2000 CREs per module on average). We then aggregated CRE modules into 13 groups with multiple steps: (1) defining ubiquitous score as the proportion of samples with average CRE activity ≥0.2 for each CRE module; (2) defining CRE modules as Broadly-active (G1) if their ubiquitous scores were above 0.5; (3) defining CRE modules as Multi-tissue (G2) if their ubiquitous scores were between 0.1 and 0.5; (4) clustering 242 reference samples into 11 clusters based on average CRE signals across 84 modules by hierarchical clustering with 1-Pearson correlation coefficient as the distance metric, naming each sample cluster manually based on the names of majority samples; and (5) grouping the rest of the CRE modules based on the sample cluster where the CRE module shows the strongest signal. The CRE modules were visualized by R package *ComplexHeatmap* (v2.4.3, R-4.0)^126^ in **Fig. 1c**.

We annotated CRE modules based on different functional categories: (1) genomic regions: annotation calculated by R package *annotatr* (v1.6.0)^127^ (**Fig. 1c**); (2) TF binding sites: we used position weight matrix (PWM) of binding sites for TF families collected in EpiMap and scanned ARE modules for enriched motifs by Homer (v4.11.1) ^40^ with background auto-selected; we filtered TF motifs based on adjust *P*-value after Benjamini-Hochberg (BH) correction (cutoff of 0.05) and odds ratio (cutoff of 1.2); we visualized the enrichment for each TF family (mean odds ratio) in promoter and enhancers (**Extended Data Fig. 1b-c**); and (3) biological processes from gene ontology (GO): enrichment of nearby genes is calculated by R package rGREAT (v1.2.1)^128^ (**Extended Data Fig. 1d**).

### Cell-type fraction estimation

We estimated fractions of six major white cell types, including CD4^+^ T cells, CD8^+^ T cells, B cells, natural killer cells, monocytes, and neutrophils in our bulk blood samples based on CRE activity from reference cell-sorted profiles. We carried out the analysis for each of three histone marks with strong cell type-specific signals (H3K27ac, H3K4me1, and H3K36me3) in four steps: (1) normalization of CRE activity including reference cell-sorted samples from Roadmap^29^ and BLUEPRINT^59^ and our samples described as above; (2) identification of cell-type marker CREs based on activity across reference samples, and then filtering marker CREs for each cell type based on correlation among each other in our bulk dataset, a strategy applied in a previous report^129^; (3) randomly sampling the same number of marker CREs (N=30 for H3K27ac and H3K4me1, and 15 for H3K36me3) from the whole marker CRE set for each cell type for 100 times and estimating cell fractions for each of our bulk samples based on activity of marker CREs from each sampling with non-negative Poisson regression in R package addreg (v3.0)^130^: 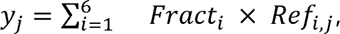 where *y_j_* denotes activity in a bulk sample for *j-th* CRE, Fract_i_ denotes the cell fraction of the bulk sample for *i-th* cell type such that *Fract_i_* ≥ 0, and *Ref_i,j_* denotes activity of *j-th* CRE in *i-th* cell type; (4) calculation of the average of resultant cell fractions estimated from each sampling as the final estimation.

### Differential CRE analysis and motif enrichment

To identify dCREs associated with BD, we applied surrogate variable analysis with R package sva (v3.28.0)^131^ followed by a linear regression model with R package limma (v.3.36.5)^132^, to account for the hidden variables (the inferred surrogate variables). The hidden variables could explain a significant proportion of the variance of residuals after regressing out the known variables, including gender, age, and disease status. We found the inferred surrogate variable was strongly correlated to the estimated cell fractions. We finally defined BD dCREs as those passing the adjusted *P*-value cutoff (0.05) after BH correction. We used the position weight matrix (PWM) of binding sites for TF families collected in EpiMap, scanned dCRE for each histone mark for enriched motifs by Homer (v4.11.1)^40^ with background auto-selected, and filtered TF motifs based on adjusted *P*-value (cutoff of 0.05) and odds ratio (cutoff of 1.2).

To identify CREs associated with drug response, we included drugs taken in at least five individuals and used the same strategy as above while including other drugs taken as covariates. We finally defined dCREs associated with drug response as those passing adjusted *P*-value cutoff (0.2) after BH correction.

### Genes with BD differential epigenomic signatures

We identified genes with BD differential epigenomic signatures based on the BD differential signal for each histone mark on the gene body and gene regulatory domain of a gene. We extracted information about the gene body from GENCODE (v26)^133^. We defined gene regulatory domain as CREs linking to TSS of a gene based on any chromatin loops inferred from promoter-capture Hi-C data of sorted circulating immune cell types from BLUEPRINT^57^, and filter any CREs overlapping with the gene body. We then aggregated differential signals of CREs over gene body and gene regulatory domain for each gene for each histone mark based on meta-analysis implemented in R package meta (v4.9-5) into z scores. We flipped the sign of z scores related to H3K27me3 since it is a repressive mark. With gene body H3K36me3 well accepted as a transcription signature, we found other scores including gene body H3K27ac, gene body H3K27me3, gene regulatory domain H3K27ac, and gene regulatory domain H3K36me3 to be strongly correlated with it. We thus included all five scores to define the genes with BD differential epigenomic signature with the following criteria: (1) at least one score passes the adjusted *P*-value cutoff (BH correction, 0.05); (2) the score for gene body H3K36me3 passes the permissive nominal *P* cutoff of 0.01; (3) at least two scores pass the nominal *P* cutoff of 0.01 and they should have a consistent sign of differential signal.

### hQTL mapping

We adapted the pipeline of our previous eGTEx study^56^ for histone QTL mapping. Briefly, we inferred the latent factors in our samples for each histone mark by Peer (R package v1.0)^134^, and identified the optimum number of Peer factors (10 for all marks) to correct for. We confirmed that QC metrics such as fraction of reads in peaks (FRiP) and RSC as well as estimated cell fractions are highly correlated with these latent factors, thus these were taken into account during QTL calling (**Extended data Fig. 3a**). We also added age and gender as covariates. We applied *FastQTL* (v2.184)^135^ to map hQTLs within 100kb to the center of each CRE on autosomes for common variants with minor allele frequency ≥ 0.05. We applied a two-step multiple test correction: (1) first estimated an empirical *P*-value for each CRE based on the lead nominal *P*-value and permutation results (--permute 1000 10000) by fitting a beta distribution to account for multiple variants tested with *FastQTL*, and (2) calculated q-value for the empirical *P*-value to account for multiple CREs tested with R package *qvalue* (v2.12.0), defined gCREs based q-value cutoff of 0.05, and defined hQTL for each CRE with corresponding nominal *P*-value threshold to the q-value cutoff 0.05.

### hQTL sharing across histone marks and between the blood and brain

We followed the strategy used in our eGTEx studies^48^ to quantify QTL sharing between two histone marks in our blood data. Briefly, for each pair of discovery mark and replication mark, we checked the lead hQTL signal for each gCRE based on the discovery mark for the replication mark and showed patterns of increasing directionality consistency between both marks with stronger nominal *P*-value signal (**Extended data Fig. 3c**). We simply assumed that non-shared QTL events lead to a 50%-50% chance to show consistent sign or not between marks, thus we estimated the proportion of overall sharing as the proportion of directionality inconsistency deducted from that of directionality consistency between two marks (**Extended data Fig. 3d**). We applied the same strategy to calculating the sharing of H3K27ac between blood and brain, with a permissive q-value cutoff of 0.2 to define gCREs in the blood (**Extended data Fig. 3f)**.

### GWAS-hQTL, hQTL-eQTL, and GWAS-eQTL colocalization and Mendelian randomization (MR)

Since the Mayo Clinic cohort is part of the Psychiatric Genomics Consortium (PGC) BD GWAS study^6^, we regenerated and used BD GWAS without the Mayoc Clinic cohort for all the downstream analyses. We applied a permissive q-value cutoff of 0.2 to define gCREs for colocalization analysis and Mendelian randomization. For both analyses, we only included gCREs within 100kb of any GWAS signal passing subthreshold cutoff (*P* ≤1×10^-6^) or eQTL signal (defined in GTEx). We used coloc (v3.2-1)^50^, a Bayesian framework, to estimate the pairwise posterior probability (PP4) of causal SNPs colocalized among GWAS, haQTL, and eQTL signals based on summary statistics. We applied Mendelian Randomization, specifically MR-Egger implemented in R package MendelianRandomization (v0.4.1)^136^, to infer the causal effect of CRE activity on either gene expression or phenotypic risk based on haQTL/eQTL/GWAS summary statistics. We filtered SNPs requiring its hQTL effect (adjusted hQTL *P*-value cutoff of 0.2, by BH test carried out for all SNPs in the locus) and pruned by r^2^ (cutoff 0.6) estimated from our cohort by PLINK (v1.9)^137^. We also summarized the BD GWAS loci that could be associated with our gCREs based on GWAS-haQTL colocalization. We clumped BD GWAS SNPs (*P* ≤1×10^-6^) with PLINK (v1.9, --clump-r2=0.2, --clump-kb=250) into 203 independent loci and identified 97 of them could be associated with the colocalized gCREs above through hQTL (q-value cutoff of 0.2) (**Supplementary Table 4**).

### Gene regulatory domain and BD driver genes prioritization

We applied two strategies to define the gene regulatory domain, or CREs linking to a specific gene: (1) physical interaction-based evidence: we aggregated chromatin loops from promoter-capture Hi-C data across multiple immune cell types from BLUEPRINT^57^ and linked a CRE to a gene if its connection to any TSS of the gene was supported by chromatin loops; and (2) genetics-based evidence: we included four of gLink scores defined in our eGTEx study^56^, (a) gARE-dist-to-FMeQTL: the shortest distance between FM-eQTLs of an eGene to a specific gCRE; (b) coloc-PP4: the coloc PP4 between hQTL of a gCRE and eQTL of an eGene; (c) coloc-PP4/3: the ratio of PP4 to PP3; (d) MR: adjusted *P*-value (BH) for MR-Egger test with hQTL and eQTL summary statistics.

We defined all the gCREs within 100kb of eGene TSS as candidate CREs and evaluated gLink scores’s performance to predict physical-interaction-based links with AUPRC, finding that top gLink scores enriched positive links. We also permuted gLink scores within each CRE-gene distance bin for 50 times and found the original gLink scores worked significantly better than the permuted gLink scores. Both results indicated the two CRE-gene linking strategies above were consistent with each other, and thus were used for the downstream analysis to define driver genes.

We defined BD driver genes as genes that are susceptible to upstream genetic signals and impact downstream changes during BD pathogenesis. Thus, we prioritized those genes whose gene regulatory domain—derived from either promoter-capture Hi-C data or gLink scores—included both gCREs sharing genetic signal with BD based on colocalization (*coloc* PP4 cutoff 0.5) or MR analysis (nominal *P* cutoff 0.01), a sign of acting in the upstream genetic susceptibility, and dCREs acting downstream of BD (adjusted *P* cutoff 0.05, BH correction).

### Latent factor analysis and annotation

We filtered dCREs with robust signals based on chromatin loops for downstream latent factor analysis. We first defined Hi-C blocks as groups of genomic regions linking with each other through chromatin loops no longer than 1Mb. We found that as the density of links in the Hi-C blocks increased, the enrichment of differential peaks also increased (**Extended Data Fig. 4a**). The enrichment pattern is quite consistent across marks and also likely to be supported by BD GWAS signal (**Extended Data Fig. 4b**). We thus only focused on dCREs (nominal *P* cutoff 0.01) in the Hi-C blocks show significant enrichment (Fisher’s exact test, BH correction, adjusted *P* cutoff 0.1) as robust differential signals, and inferred latent factors across all individuals from all five marks with R package MOFA (v1.6.2, numFactors =40, dropFactorThreshold =0.005). We then annotated these factors by revealing the significant correlation (adjust *P* cutoff 0.05, BH correction across all terms, R^2^ cutoff 0.1) of latent factors with EHR and cell fraction estimated across BD patients (**Extended Data Fig. 5c**).

### Patient subtyping, annotation and marker CREs

We calculated the Pearson’s correlation coefficients across latent factors among all individuals and clustered participants into five groups based on the correlation matrix with hierarchical clustering (R function *hclust*, **Fig. 5a**). We next identified EHR terms enriched in each patient group compared to the rest of patients (Fisher’s exact test for categorical terms, *t*-test for numerical terms, nominal *P* cutoff 0.02, **Fig. 5c**), and visualized it with R package ComplexHeatmap (v2.4.3, R-4.0)^126^.

We only focused on the dCREs with robust signal above (nominal *P* cutoff 0.01, and within Hi-C blocks enriched for dCREs, adjusted *P* cutoff 0.1) for marker CREs detection. We applied linear regression to detect marker CREs for each patient group compared to the rest of the patients with gender and age as covariates (adjusted *P* cutoff 0.05, BH correction). We then annotated marker CREs with R package rGREAT (v1.2.1)^128^ (Fisher’ exact test, adjusted *P* cutoff 0.05, BH correction) and visualized it with R package pheatmap (v1.0.12)^138^.

### Epi-partitioned PRSs

We followed a standard PRS pipeline of clumping to build the full PRS model^139^ based on BD GWAS summary statistics. During the quality control step of GWAS summary statistics, we filter SNPs based on their proportions of the presented individuals (call rate≥0.9), minor allele frequency (MAF≥0.01), and imputation quality score (INFO≥0.8). We then carried out clumping by PLINK (v1.9)^137^ with parameters of linkage disequilibrium (LD) R^2^ cutoff (0.4) and distance cutoff (100kb) based on the LD estimated from the Mayo Clinic cohort. We finally generated PRSs by aggregating effects of all selected SNPs for each genotyped individual by PLINK.

To generate epi-partitioned PRSs, we first collected various CRE sets (**Extended Data Fig. 5e**), including those blood CRE groups showing co-activity patterns across reference epigenomes in our study (**Fig. 1b**), CRE groups detected in four tissues from our previous eGTEx report^56^, BD differential CREs in blood, blood gCREs, and the blood gCREs sharing BD genetics based on two statistical methods coloc and MR. Next, for each epi-partitioned PRS, we selected the SNPs located near the corresponding CRE set by bedtools (bedtools window -w 2000) before clumping with the same parameters as the full model.

### Drug repurposing analysis

We downloaded 49,488 differential gene expression profiles of 12k genes from hematopoietic lineages in response to 33,627 compounds from CMap (LINCS 2020). We then tested in these profiles the enrichment of gene sets with BD up-/down-regulated epigenomic signatures identified in a previous step (**Fig. 2b-c**) by Gene Set Enrichment Analysis (GSEA)^84^ implemented in the Python package GSEApy^140^ and run with the following parameters: permutation_num=1000, max_size=2000. We only considered drugs/compounds “BD-repurposed” if their corresponding differential expression profiles (1) significantly enriched both BD gene sets (*P*<0.001, the minimum *P*-value could be achieved with 1000 times of permutations) by GSEA, (2) showed down-regulation of BD up-regulated genes, and (3) showed up-regulation of BD down-regulated genes. Drug annotation with ATC code in **Fig. 6c** and **Extended Data Fig. 6d-e** was downloaded from DrugBank v5.1.12^141^.

